# Prediction of mild cognitive impairment progression using time-sensitive multimodal biomarkers

**DOI:** 10.1101/2025.09.20.25336240

**Authors:** Jonathan Gallego-Rudolf, Alex I. Wiesman, Yara Yakoub, Henrik Zetterberg, Kaj Blennow, Sylvain Baillet, Sylvia Villeneuve, the PREVENT-AD Research Group

**Author notes:** Corresponding Authors. Sylvia Villeneuve, PhD. Douglas Research Centre Department of Psychiatry McGill University, Montreal, QC, Canada.

## Abstract

Alzheimer’s disease (AD) develops over a prolonged asymptomatic phase marked by silent pathology. Identifying cognitively unimpaired individuals likely to progress to mild cognitive impairment (MCI) is essential for early intervention. We investigated whether multimodal combinations of biomarkers, including frequency-specific neurophysiological activity, enhance prediction beyond demographic and genetic factors in older adults (n = 102; 31 progressors; mean follow-up = 5.9 years). Biomarkers included MEG-derived alpha power, MRI-derived hippocampal volume, plasma Aβ42/40 ratio and p-tau217, and neocortical Aβ and entorhinal tau PET. Cox regression models estimated progression risk and tested time-varying prognostic effects. Neurophysiological and proteinopathy biomarkers improved prediction beyond clinical and genetic factors. Elevated alpha power predicted short-term risk, but its predictive value weakened over time, whereas high neocortical Aβ became increasingly predictive with longer follow-up. Plasma Aβ42/40, p-tau217, and tau PET each conferred higher risk, while hippocampal volume did not. Findings support a multimodal, time-sensitive framework for individualized risk prediction in preclinical AD.

## Main

Alzheimer’s disease (AD) progresses along a prolonged continuum, beginning with an asymptomatic phase during which neuropathological changes develop silently (Dubois et al., 2016; Jack et al., 2013, 2024; Sperling et al., 2011). Mild cognitive impairment (MCI) marks the prodromal stage of the disease, with measurable deficits on neuropsychological testing that typically do not interfere with daily functioning (Petersen et al., 1999; Petersen & Morris, 2005). Identifying cognitively unimpaired individuals at higher risk of progression to MCI is critical for early intervention and clinical trial targeting (Aisen et al., 2022; Jack et al., 2024).

Biologically, AD is characterized by the pathological deposition of amyloid-beta (Aβ) into plaques and hyperphosphorylated tau protein into tangles and dystrophic neurites in the brain (Alzheimer, 1907; Braak & Braak, 1991, 1995; Glenner & Wong, 1984; Jack et al., 2024). These proteinopathies emerge many years before the onset of cognitive symptoms (Jack et al., 2010, 2024) and trigger a cascade of pathological processes that ultimately lead to neurodegeneration and cognitive decline (Aschenbrenner et al., 2018; Bloom, 2014; Busche & Hyman, 2020; Gulisano et al., 2018). Current biological criteria for AD emphasize in-vivo quantification of Aβ and tau using positron emission tomography (PET) and blood-based biomarkers, such as soluble Aβ species (e.g., Aβ42, Aβ40) and their Aβ42/40 ratio, and phosphorylated tau protein at threonine 217 (p-tau217), as core diagnostic tools. Additional biomarkers derived from magnetic resonance imaging (MRI) and fluorodeoxyglucose PET, along with plasma and cerebrospinal fluid (CSF) markers of specific processes involved in AD pathophysiology (i.e., tau phosphorylation status, inflammation, and grey matter neurodegeneration) have also been proposed to stage disease severity and identify co-pathologies (Jack et al., 2024).

While proteinopathy biomarkers predict progression from normal cognition to MCI (Ossenkoppele et al., 2025; Schindler et al., 2021; Soldan et al., 2025; Yakoub et al., 2025), most studies have assessed them in isolation, without integrating measures of brain function (Gonneaud et al., 2021; López-Sanz et al., 2018) or evaluating whether their prognostic value changes over time.

Functional neurophysiology, measured noninvasively with electroencephalography (EEG; Babiloni et al., 2021) or magnetoencephalography (MEG; Baillet, 2017; López-Sanz et al., 2018), can detect alterations in brain activity linked to Aβ and tau deposition. In symptomatic AD, neurophysiological slowing, with increased low-frequency (delta-theta; 1–8 Hz) and decreased higher-frequency (alpha; 8–12 Hz) activity, is closely linked to Aβ and tau deposition (Ranasinghe et al., 2020; Wiesman et al., 2022). In the asymptomatic stage, a nuanced pattern emerges (Nakamura et al., 2017, 2018). We recently showed that early Aβ accumulation is associated with enhanced alpha activity, whereas rising subsequent tau pathology in the temporal lobe corresponds to a shift toward slower brain activity (Gallego-Rudolf et al., 2024). These patterns suggest that neurophysiological dynamics may provide complementary, stage-dependent prognostic information to proteinopathy measures, offering a functional perspective on disease stage that could improve time-sensitive risk prediction.

Here, we test whether integrating functional neurophysiology with established proteinopathy and structural biomarkers improves prediction of progression to MCI in asymptomatic older adults at elevated familial risk of AD. Using data from the Pre-Symptomatic Evaluation of Experimental or Novel Treatment for Alzheimer’s Disease cohort (PREVENT-AD; Breitner et al., 2016), we combined MEG-derived alpha-band activity, MRI-derived hippocampal volume, plasma and PET measures of Aβ and tau, and demographic and genetic information in Cox regression models to estimate individual risk of progression (**Figure 1**). We further modeled time-varying effects to determine whether the prognostic value of each biomarker changed over time. This approach allowed us to evaluate both the additive value of functional brain measures and the stage-dependent trajectories of multiple biomarkers, advancing a multimodal, time-sensitive framework for preclinical AD risk prediction.

**Figure 1.**
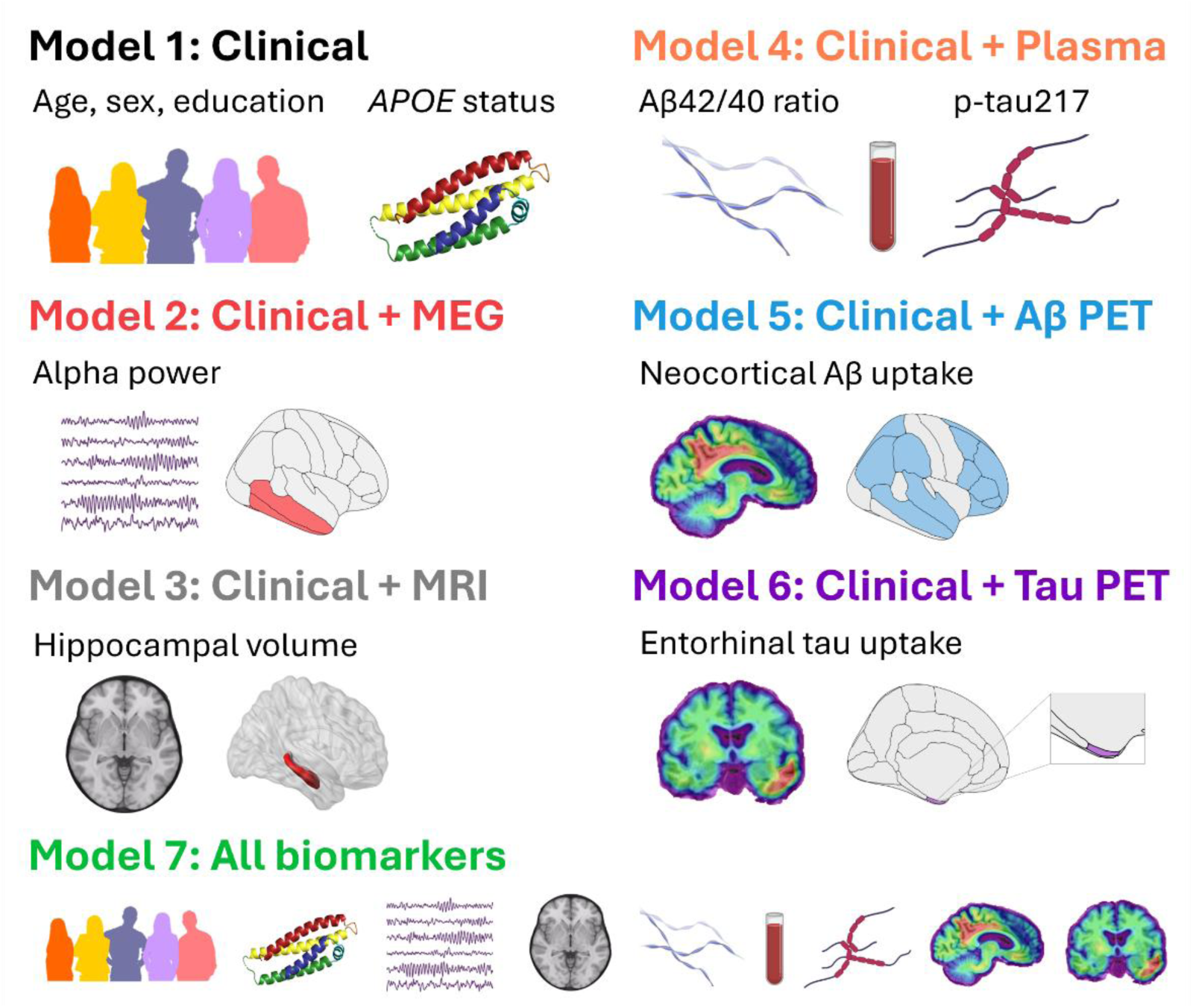
Cox proportional hazard models for predicting MCI progression. A series of Cox proportional hazard models assessed the added value of incorporating neurophysiological, brain structural, and proteinopathy biomarkers for estimating the risk of progression to MCI. The reference clinical model (Model 1) included only demographic and genetic variables (age, sex, education, and *APOE* ε4 status). Subsequent models added: alpha-band relative power derived from MEG source time series mapped to temporal cortical regions (Model 2); MRI-derived hippocampal volume normalized to the total intracranial volume (Model 3); plasma Aβ42/40 ratio and p-tau217 (Model 4); neocortical Aβ PET uptake (Model 5); or entorhinal tau PET uptake (Model 6). The final model (Model 7) combined clinical information with all available biomarkers. Brain schematics were adapted using the *ggseg* package in *R*; other graphical elements were created using *Biorender*.

## Results

### Participants characteristics

All participants were asymptomatic at the time of biomarker collection, based on a multidisciplinary clinical consensus review (see *Neuropsychological assessment and MCI diagnosis* section in the Methods). Over a mean follow-up of 5.88 years (SD = 1.64; range = 0.19–7.45), 31 participants progressed to MCI and 71 remained stable. Compared with non-progressors, progressors were significantly older (*W* = 748.5; *p*_(FDR)_ = 0.0211), had lower plasma Aβ42/40 ratios (*W* = 1501; *p*_(FDR)_ = 0.0090), and showed higher plasma p-tau217 concentrations (*W* = 639; *p*_(FDR)_ = 0.0026). They also exhibited greater neocortical Aβ PET uptake (*W* = 473; *p*_(FDR)_ < 0.00001) and entorhinal tau PET uptake (*W* = 577; *p*_(FDR)_ = 0.0007; **Table 1**).

**Table 1.**
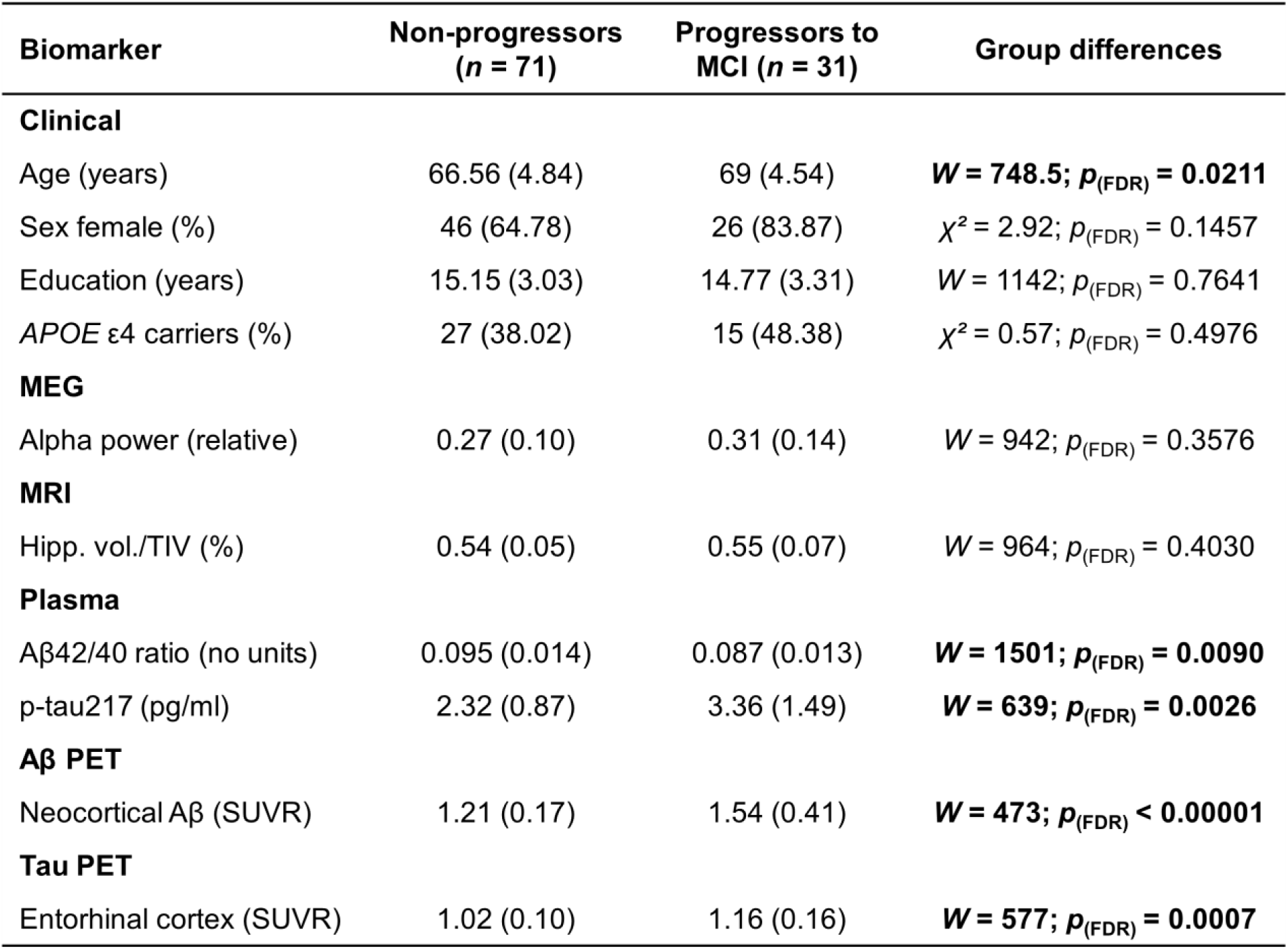
Characteristics of PREVENT-AD participants. Comparison of clinical, neuroimaging, and plasma biomarkers between individuals who remained cognitively unimpaired (non-progressors, *n* = 71) and individuals who developed MCI (progressors, *n* = 31). All variables are presented as group means (standard deviations), except for sex and *APOE* ε4 carrier status, which are reported as counts (percentages). Group differences were assessed using Wilcoxon rank-sum tests for continuous variables and chi-square tests for categorical variables. Statistical significance was set at *p* < 0.05, and significant p-values are shown in boldface. Progressors were, on average, older, had lower plasma Aβ42/40 ratio, and exhibited higher plasma p-tau217 concentrations, neocortical Aβ PET uptake, and entorhinal tau PET uptake. Hipp. vol., hippocampal volume; pg/ml, picograms per milliliter; SUVR, standardized uptake value ratio; TIV, total intracranial volume.

| Biomarker | Non-progressors<br>($n = 71$) | Progressors to<br>MCI ($n = 31$) | Group differences |
| --- | --- | --- | --- |
| **Clinical** |  |  |  |
| Age (years) | 66.56 (4.84) | 69 (4.54) | **$W = 748.5$; $p_{(FDR)} = 0.0211$** |
| Sex female (%) | 46 (64.78) | 26 (83.87) | $\chi^2 = 2.92$; $p_{(FDR)} = 0.1457$ |
| Education (years) | 15.15 (3.03) | 14.77 (3.31) | $W = 1142$; $p_{(FDR)} = 0.7641$ |
| APOE $\epsilon 4$ carriers (%) | 27 (38.02) | 15 (48.38) | $\chi^2 = 0.57$; $p_{(FDR)} = 0.4976$ |
| **MEG** |  |  |  |
| Alpha power (relative) | 0.27 (0.10) | 0.31 (0.14) | $W = 942$; $p_{(FDR)} = 0.3576$ |
| **MRI** |  |  |  |
| Hipp. vol./TIV (%) | 0.54 (0.05) | 0.55 (0.07) | $W = 964$; $p_{(FDR)} = 0.4030$ |
| **Plasma** |  |  |  |
| Aβ42/40 ratio (no units) | 0.095 (0.014) | 0.087 (0.013) | **$W = 1501$; $p_{(FDR)} = 0.0090$** |
| p-tau217 (pg/ml) | 2.32 (0.87) | 3.36 (1.49) | **$W = 639$; $p_{(FDR)} = 0.0026$** |
| **Aβ PET** |  |  |  |
| Neocortical Aβ (SUVR) | 1.21 (0.17) | 1.54 (0.41) | **$W = 473$; $p_{(FDR)} < 0.00001$** |
| **Tau PET** |  |  |  |
| Entorhinal cortex (SUVR) | 1.02 (0.10) | 1.16 (0.16) | **$W = 577$; $p_{(FDR)} = 0.0007$** |

For broader context, we also examined three extended PREVENT-AD samples comprising the main analysis cohort plus participants with available plasma or PET biomarkers but no MEG scans. These included n = 211 with plasma data (**Extended Table 1**), n = 226 with Aβ PET (**Extended Table 2**) and n = 224 with tau PET (**Extended Table 3**).

### Neurophysiology and proteinopathy biomarkers predict MCI progression

In the main cohort (n = 102), Cox proportional hazard models tested whether the biomarker measurements—MEG alpha-band relative power, MRI-derived hippocampal volume, plasma Aβ42/40 ratio and p-tau217, neocortical Aβ PET, and entorhinal tau PET—could improve prediction beyond a reference clinical model of age, sex, years of education, and *APOE* ε4 status (**Figure 2**; **Figure 3**).

**Figure 2.**
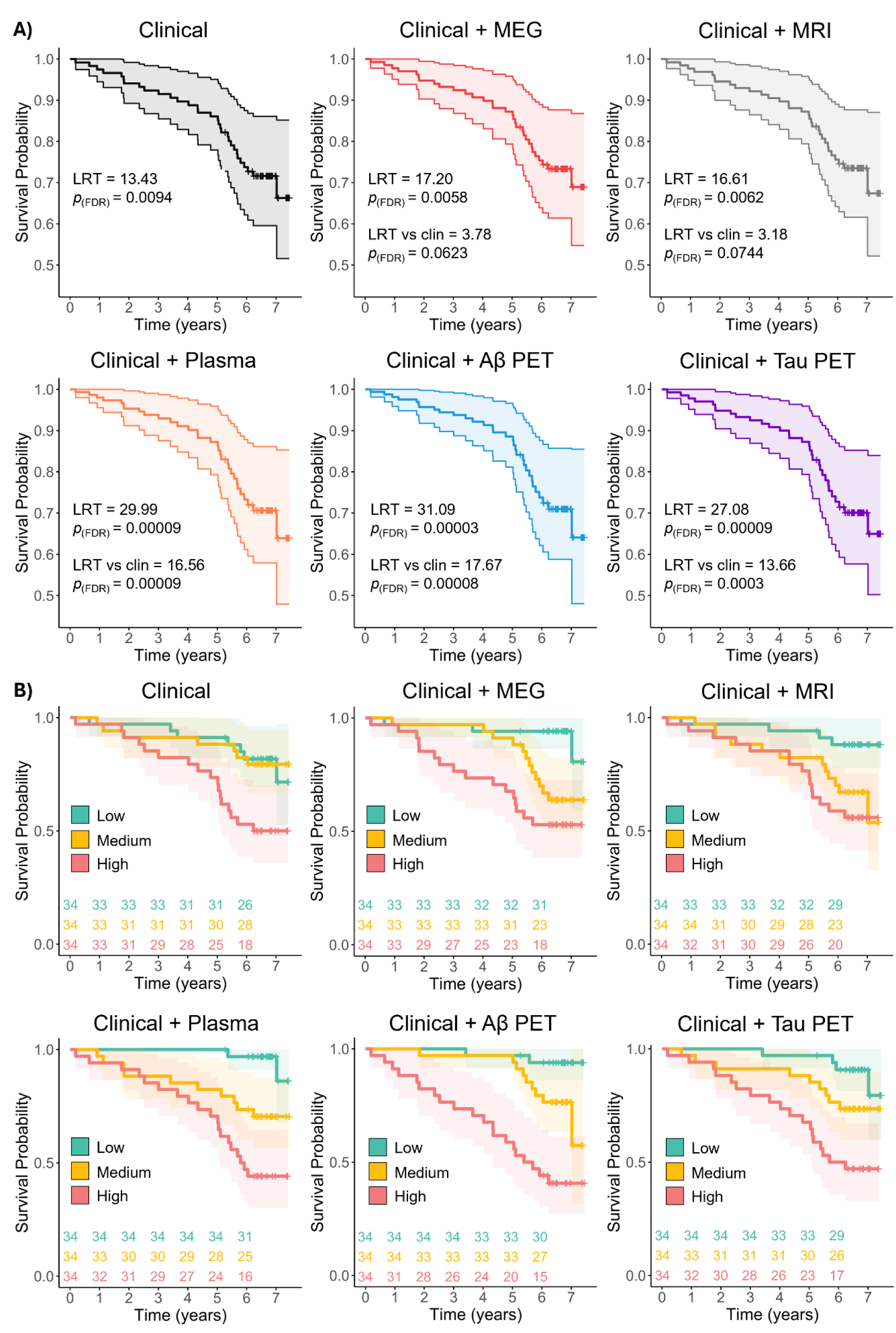
Cox regression survival analysis. **A)** Survival curves for the reference clinical model and for each model combining clinical information with neurophysiological activity, hippocampal volume and proteinopathy biomarkers. Likelihood ratio tests (LRTs) indicated that all models significantly outperformed the null model. Pairwise comparisons with the clinical model (LRT vs clin) showed that adding plasma biomarkers, neocortical Aβ PET or entorhinal tau PET significantly improved predictive performance. P-values were corrected for multiple comparisons using the false discovery rate (FDR). Thick lines represent the estimated survival curves for the full sample; shaded areas show 95% confidence intervals (CIs). **B)** Participants were stratified into low-, medium-, and high-risk tertiles based on risk scores derived from each model. Survival curves are shown to illustrate the discriminative accuracy of the models in predicting progression to MCI. As in panel A, thick lines represent the estimated survival curves for each risk group, and shaded areas indicate 95% CIs. The number of participants remaining cognitively stable at each follow-up year is shown below each panel.

**Figure 3.**
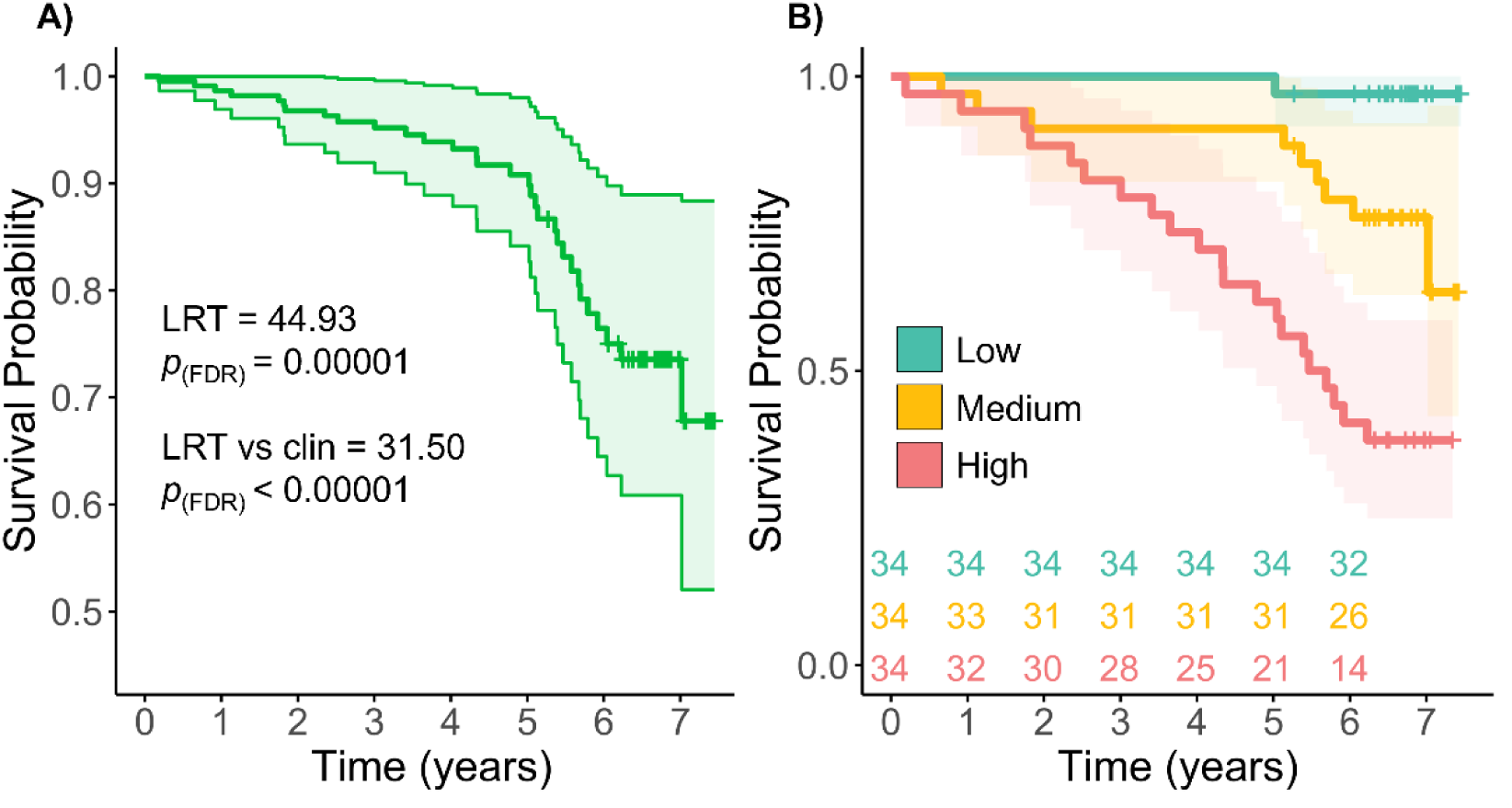
Cox regression survival analysis including all biomarkers. **A)** Survival curve for the model combining clinical information with neurophysiological activity, hippocampal volume, and proteinopathy biomarkers. Likelihood ratio tests (LRTs) indicated that this model significantly outperformed both the null and reference clinical models. P-values were corrected for multiple comparisons using the false discovery rate (FDR). The thick line represents the estimated survival curve for the full sample; shaded area shows 95% confidence intervals (CIs). **B)** Participants were stratified into low-, medium-, and high-risk tertiles based on the risk scores derived from the all-biomarker model. Survival curves are shown for each group to illustrate the discriminative accuracy of the model in predicting progression to MCI. As in panel A, thick lines represent the estimated survival curves for each risk group, and shaded areas indicate 95% CIs. The number of participants remaining cognitively stable at each follow-up year is shown below each panel.

All models significantly outperformed the null model. The clinical model alone was predictive (likelihood ratio test, LRT = 13.43, *p*_(FDR)_ = 0.0094). Adding MEG alpha power (LRT = 17.20, *p*_(FDR)_ = 0.0058), hippocampal volume (LRT = 16.61, *p*_(FDR)_ = 0.0062), plasma biomarkers (LRT = 29.99, *p*_(FDR)_ = 0.00009), Aβ PET (LRT = 31.09, *p*_(FDR)_ = 0.00003), or entorhinal tau PET (LRT = 27.08, *p*_(FDR)_ = 0.00009) each improved model fit (**Figure 2A**). A full multimodal model including all biomarkers yielded the strongest performance (LRT = 44.93, *p*_(FDR)_ < 0.00001; **Figure 3A**).

When compared directly against the reference clinical model, plasma biomarkers (LRT = 16.56, *p*_(FDR)_ = 0.00009), Aβ PET (LRT = 17.67, *p*_(FDR)_ = 0.00008), and tau PET (LRT = 13.66, *p*_(FDR)_ = 0.0003) significantly improved prediction after correcting for multiple comparisons (**Figure 2A**). By contrast, the effects of MEG alpha power (LRT = 3.78; *p*_(FDR)_ = 0.0623) and hippocampal volume (LRT = 3.18, *p*_(FDR)_ = 0.0744) did not survive correction. Findings were consistent using the Akaike Information Criterion (AIC) and concordance index metrics (**Supplementary Figure 1**).

The prognostic value of proteinopathy biomarkers replicated in the extended samples: plasma (*n* = 211; LRT = 35.51, *p*_(FDR)_ < 0.00001; LRT vs clinical = 14.44, *p*_(FDR)_ = 0.0001), Aβ PET (*n* = 226; LRT = 34.95, *p*_(FDR)_ < 0.00001; LRT vs clinical = 17.65, *p*_(FDR)_ = 0.00004), and tau PET (*n* = 224; LRT = 35.16, *p*_(FDR)_ < 0.00001; LRT vs clinical = 18.11, *p*_(FDR)_ = 0.00004; **Supplementary Figure 2A**).

Risk-stratified Kaplan-Meier curves showed better separation between low-, medium-and high-risk tertiles when biomarker data were included (**Figure 2B**, **Figure 3B**). Similar patterns were observed in the extended replication analyses (**Supplementary Figure 2B**).

In the full multimodal model, higher risk scores were associated with older age (*t* = 3.47, *p*_(FDR)_ = 0.0011), female sex (*t* = -4.48, *p*_(FDR)_ = 0.00008), being an *APOE* ε4 allele carrier (*t* = -2.21, *p*_(FDR)_ = 0.0366), higher MEG alpha power (*t* = 3.52, *p*_(FDR)_ = 0.0011), lower plasma Aβ42/40 ratio (*t* = -4.06, *p*_(FDR)_ = 0.0002), higher plasma p-tau217 (*t* = 6.61, *p*_(FDR)_ < 0.00001), greater Aβ PET burden (*t* = 8.18, *p*_(FDR)_ < 0.00001), and higher entorhinal tau PET uptake (*t* = 10.21, *p*_(FDR)_ < 0.00001; **Figure 4**). Years of education (*t* = 0.57, *p*_(FDR)_ = 0.5670) and hippocampal volume (*t* = 1.82, *p*_(FDR)_ = 0.0794) were not significant predictors. Associations remained consistent in biomarker-specific models (**Supplementary Table 4**) and were not attributable to multicollinearity (**Supplementary Figure 3**).

**Figure 4.**
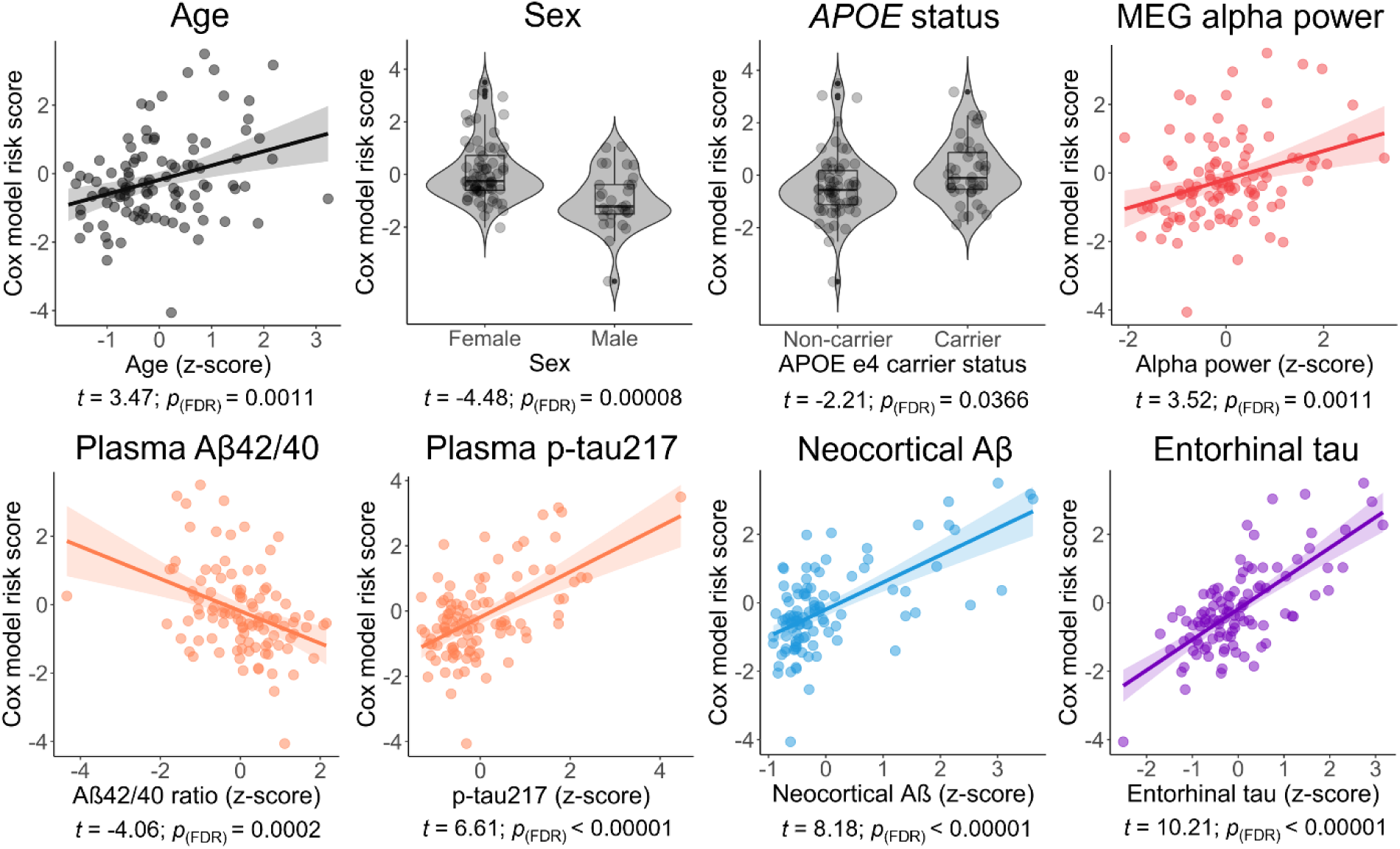
Biomarker associations with Cox proportional-hazard risk scores. Scatterplots and violin/boxplots show the relationship between individual biomarker values and participants’ risk scores derived from the Cox regression model including all biomarkers. Associations with continuous variables were assessed using linear regression. Risk scores were positively associated with age, MEG alpha-band power, plasma p-tau217, neocortical Aβ PET uptake, and entorhinal tau PET uptake, and negatively associated with plasma Aβ42/40 ratio. Education and hippocampal volume were not significantly associated with risk scores and are not shown. Regression lines indicate the fitted linear trend; shaded areas denote 95% confidence intervals (CIs). Sex and *APOE* ε4 carrier status were treated as categorical variables and tested using independent-sample *t*-tests. Group differences are illustrated using boxplots embedded in violin plots, showing the group median, interquartile range, and data distribution. Higher risk scores were observed in females compared to males, and in *APOE* ε4 carriers compared to non-carriers. The *t*-statistics and FDR-corrected *p*-values (adjusted for multiple comparisons) are reported below each panel. FDR, false discovery rate.

### Time-varying effects of neurophysiology and neocortical Aβ

We next tested whether the prognostic value of each biomarker changed over the follow-up period by adding biomarker × time interaction terms to the Cox regression models. In these models, the main effect represents the hazard ratio at baseline (time = 0), whereas the time interaction term captures how that effect evolves longitudinally. Incorporating time-varying effects for alpha-band power and neocortical Aβ PET significantly improved the model discrimination (**Figure 5; Supplementary Figure 1**), consistent with preliminary proportional hazards assumption violations in standard Cox models (alpha power: *χ*² = 3.67, *p* = 0.0550; neocortical Aβ PET: *χ*² = 8.19, *p* = 0.0042).

**Figure 5.**
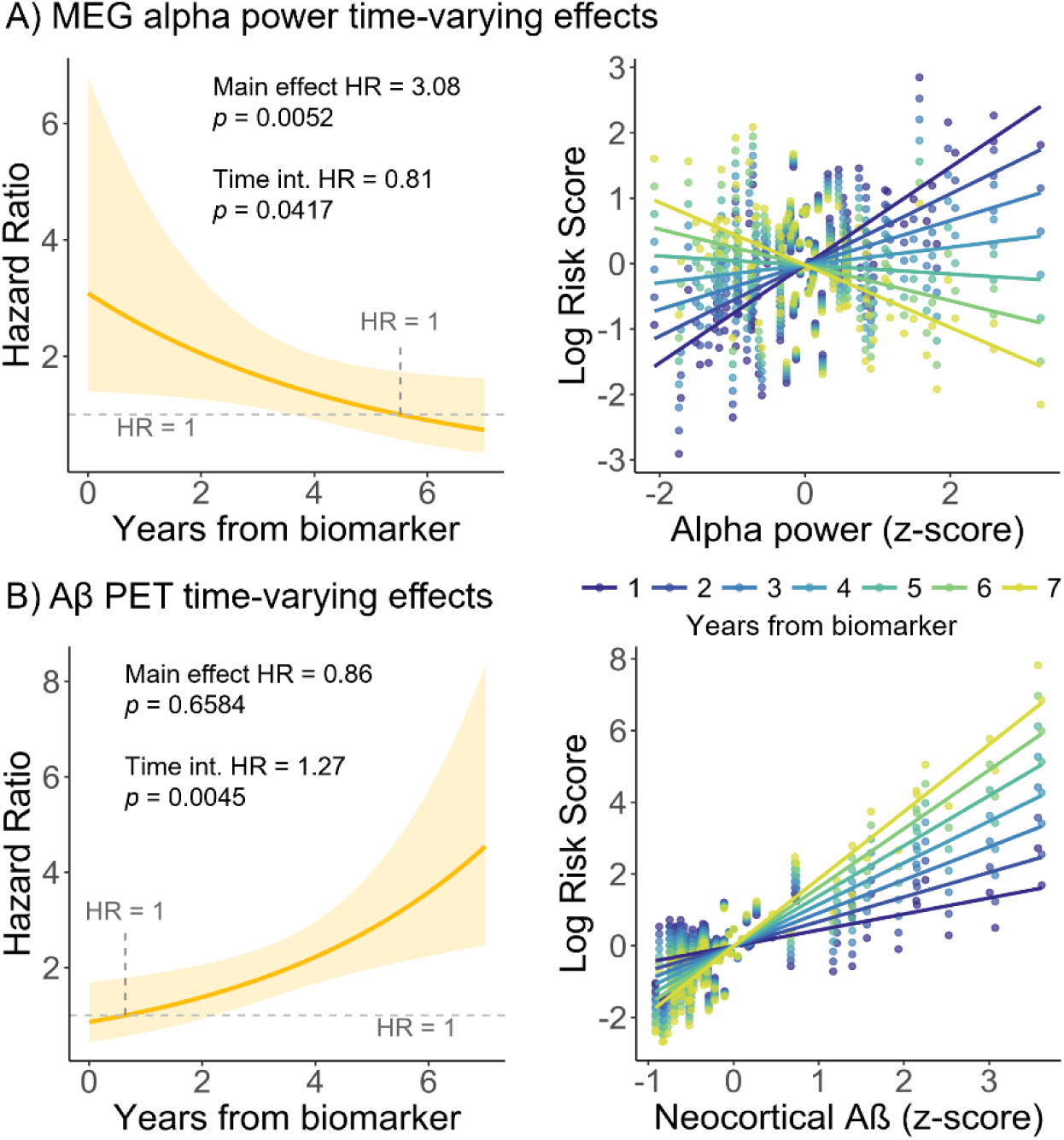
Time-varying effects of MEG alpha power and neocortical Aβ on MCI risk. We observed significant time-dependent interactions, indicating time-varying effects of alpha-band MEG activity **(A)** and neocortical Aβ PET uptake **(B)** on the risk of progression to MCI. <u>Left panels:</u> The hazard ratio is plotted over time, derived from Cox regression models that included both the main effect and time interaction terms for each biomarker. Model estimates were adjusted for age, sex, education, and *APOE* ε4 carrier status. Shaded areas represent 95% confidence intervals (CIs). The HR coefficients and associated *p*-values for the main and time interaction effects are shown in each plot. HR, hazard ratio; Time int., time interaction. <u>Right panels:</u> Time-varying associations between the z-scored biomarker values and the predicted log risk scores, with separate regression lines for each follow-up year (color-coded from dark blue to yellow, years 1–7). These plots illustrate how the relationship between each biomarker and progression risk evolves over time, while accounting for covariates. Significant time interaction terms for both MEG alpha power and Aβ PET uptake support their time-varying effects on disease progression.

Higher alpha power was associated with increased short-term risk of progression (main effect log(HR) = 1.13, HR = 3.08, *p* = 0.0052), but this association weakened over time, such that lower alpha-power became the stronger predictor later in the disease continuum (alpha × time log(HR) = -0.20, HR = 0.81, *p* = 0.0417; **Figure 5A**). In contrast, elevated neocortical Aβ PET uptake was not predictive at baseline (main effect log(HR) = -0.15, HR = 0.86, *p* = 0.6584) but its prognostic value increased steadily with time (Aβ × time log(HR) = 0.24, HR = 1.27, *p* = 0.0045; **Figure 5B**).

No significant time-dependent effects were observed for age, years of education, hippocampal volume, plasma Aβ42/40 ratio, plasma p-tau217, or entorhinal tau PET uptake (all *p* > 0.09; **Supplementary Figure 4**).

Replication in the extended Aβ PET sample confirmed the increasing prognostic value of neocortical Aβ over time (main effect log(HR) = -0.01, HR = 0.99, *p* = 0.9621; neocortical Aβ × time log(HR) = 0.18, HR = 1.20, *p* = 0.0023). In contrast, extended-sample analyses for plasma Aβ42/40 ratio, plasma p-tau217, and entorhinal tau PET again showed no significant time interactions (all *p* > 0.38; **Supplementary** Figure 5).

### Neurophysiological dynamics complement proteinopathy biomarkers

We next tested whether modeling the time-varying effects of neurophysiological activity provided predictive value beyond clinical variables and established plasma and PET-based proteinopathy biomarkers. Likelihood ratio tests showed that adding the time-dependent effect of alpha power significantly improved prediction when combined with clinical information and plasma biomarkers (LRT = 5.12, *p*_(FDR)_ = 0.0237), time-varying Aβ PET uptake (LRT = 5.60, *p*_(FDR)_ = 0.0237), or entorhinal tau PET uptake (LRT = 11.38, *p*_(FDR)_ = 0.0022).

To further evaluate the added value of neurophysiological dynamics, we used stepwise Cox regression starting from a full model that included all biomarkers plus the significant time-dependent interaction terms for alpha power and neocortical Aβ PET. The algorithm selected a final parsimonious model comprising age (log(HR) = 0.32, HR = 1.38, *p* = 0.1326), hippocampal volume (log(HR) = 0.39, HR = 1.48, *p* = 0.0661), the time-varying effect of alpha power (log(HR) = 0.01, HR = 1.01, *p* = 0.0047), neocortical Aβ PET burden (log(HR) = 0.44, HR = 1.55, *p* = 0.0086), and entorhinal tau PET uptake (log(HR) = 0.60, HR = 1.82, *p* = 0.0027) as the most informative feature set for predicting progression to MCI (**Table 2**). These results underscore the complementary role of neurophysiological dynamics in enhancing prognostic models that already include clinical information and proteinopathy biomarkers.

**Table 2.** Step-wise Cox regression. We implemented a stepwise Cox regression model selection procedure, starting from a full model that included all biomarkers and the significant time-interaction terms (t) for alpha-band neurophysiological activity and neocortical Aβ PET burden. Predictors were iteratively removed based on minimizing the Akaike Information Criterion (AIC), yielding a parsimonious model that retained predictive performance. The table shows the variables included at each step, the variables removed, and the resulting AIC value. The final model retained age, hippocampal volume, the time-dependent effect of alpha power, neocortical Aβ PET uptake, and entorhinal tau PET uptake as the best combination of predictors of progression to MCI, with only the neurophysiological and proteinopathy markers emerging as significant predictors in the final model. AIC, Akaike information criterion; Hipp. vol., hippocampal volume.

| Steps | Cox regression model | Removed | Model AIC |
| --- | --- | --- | --- |
| **Step 1** | Age, Sex, Education, *APOE*, Alpha, Alpha (t), Hipp. vol., Aβ42/40 ratio, p-tau217, Aβ PET, Aβ PET (t), Tau PET | - | 250.21 |
| **Step 2** | Age, Sex, Education, Alpha, Alpha (t), Hipp. vol., Aβ42/40 ratio, p-tau217, Aβ PET, Aβ PET (t), Tau PET | *APOE* | 248.26 |
| **Step 3** | Age, Sex, Alpha, Alpha (t), Hipp. vol., Aβ42/40 ratio, p-tau217, Aβ PET, Aβ PET (t), Tau PET | Education | 246.33 |
| **Step 4** | Age, Sex, Alpha, Alpha (t), Hipp. vol., p-tau217, Aβ PET, Aβ PET (t), Tau PET | Aβ42/40 ratio | 244.96 |
| **Step 5** | Age, Sex, Alpha (t), Hipp. vol., p-tau217, Aβ PET, Aβ PET (t), Tau PET | Alpha | 243.85 |
| **Step 6** | Age, Sex, Alpha (t), Hipp. vol., Aβ PET, Aβ PET (t), Tau PET | p-tau217 | 242.80 |
| **Step 7** | Age, Sex, Alpha (t), Hipp. vol., Aβ PET, Tau PET | Aβ PET (t) | 242.42 |
| **Step 8** | Age, Alpha (t), Hipp. vol., Aβ PET, Tau PET | Sex | 242.33 |

## Discussion

The early identification of individuals along the AD continuum is critical for the success of therapeutic interventions aimed at preventing or slowing cognitive decline (Aisen et al., 2022; Jack et al., 2024). In this longitudinal study of asymptomatic older adults with elevated familial risk of sporadic AD, we assessed whether integrating neurophysiological activity with plasma and PET proteinopathy biomarkers could improve prediction of progression to MCI beyond demographic and genetic information. We found that adding MEG-derived alpha-band spectral power and molecular biomarkers improved predictive performance relative to a clinical model alone. Critically, the risk associated with alpha power and neocortical Aβ PET burden followed time-dependent trajectories, indicating that their prognostic value changes over the disease course. Modeling these neurophysiological dynamics conferred added predictive value beyond proteinopathy biomarkers, enhancing individualized risk stratification.

We observed a stage-dependent association between alpha-band activity and future progression to MCI. Elevated alpha power predicted higher short-term risk, consistent with early cortical hyperexcitability or compensatory network dynamics (Bruña et al., 2023; Gallego-Rudolf et al., 2024; Nakamura et al., 2018; Ranasinghe et al., 2025). This effect weakened over time, suggesting that closer to symptom onset, high alpha power no longer confers elevated risk, possibly reflecting a shift towards network breakdown (Alexandersen et al., 2023; López-Sanz et al., 2017). This biphasic trajectory aligns with our previous work in PREVENT-AD, showing that early Aβ accumulation is associated with elevated alpha activity, whereas subsequent tau pathology corresponds to slowing of cortical rhythms (Gallego-Rudolf et al., 2024; see also Alexandersen et al., 2023; López-Sanz et al., 2018). The current findings extend this framework by demonstrating that alpha-band activity is not only stage-dependent but also contributes unique, time-sensitive prognostic information beyond proteinopathy measures.

Indeed, neurophysiological changes have long been recognized as a hallmark of symptomatic AD, with converging evidence pointing to widespread alterations in cortical rhythmic activity (De Haan et al., 2008; Garcés et al., 2013; Wiesman et al., 2022). These include the slowing of oscillatory activity (typically observed as increased delta and decreased alpha-band power) and are readily measurable via MEG and EEG (Ranasinghe et al., 2020; Wiesman et al., 2021, 2022). Neurophysiological slowing has been associated with both proteinopathy burden (Ranasinghe et al., 2020, 2021; Wiesman et al., 2022) and cognitive dysfunction (Babiloni et al., 2021; López-Sanz et al., 2016; Wiesman et al., 2022).

In contrast, hippocampal volume did not significantly enhance prediction, underscoring that functional measures of network dynamics may better complement molecular biomarkers for identifying individuals at high risk of MCI within a relatively short follow-up (∼7 years).

Proteinopathy biomarkers (particularly plasma Aβ42/40 ratio, plasma p-tau217, and PET measures of neocortical Aβ and entorhinal tau) were robust predictors of progression, consistent with previous work, including studies with PREVENT-AD (Ossenkoppele et al., 2025; Schindler et al., 2021; Soldan et al., 2025; Strikwerda-Brown et al., 2022; Yakoub et al., 2023, 2025). These associations held in extended samples that doubled our analytic cohort. While PET offers superior anatomical specificity, plasma-based assays reflecting Aβ metabolism and tau phosphorylation state are more accessible, minimally invasive, and increasingly scalable in clinical settings contexts (Aisen et al., 2022; Jack et al., 2024).

The time-varying effect of neocortical Aβ PET burden supports the amyloid cascade hypothesis (Hardy & Higgins, 1992; Selkoe & Hardy, 2016). Early in preclinical disease, detectable plaques may not strongly predict short-term progression. Over time, as cortical amyloid burden enters an accelerated growth phase, it becomes a stronger risk factor (Baek et al., 2020; Jack et al., 2010; Lee et al., 2022; Mattsson et al., 2019; Thal et al., 2002), likely reflecting downstream processes such as tau spread beyond the medial temporal lobe (Sanchez et al., 2021; Van Der Kant et al., 2020; Vogel et al., 2020, 2023). Notably, plasma Aβ42/40 ratio and tau biomarkers did not show significant time-varying effects, suggesting that Aβ PET may uniquely capture a pathological transition point critical for clinical progression (Baek et al., 2020; Lee et al., 2022).

In our combined models, incorporating time-varying alpha power consistently improved predictive performance beyond proteinopathy biomarkers alone. Stepwise Cox regression retained the time-dependent alpha effect alongside neocortical Aβ PET and entorhinal tau uptake, highlighting its complementary role (Babiloni et al., 2021; Maestú et al., 2019). These findings suggest that alpha-band dynamics may index both early compensatory/excitotoxic mechanisms and later-stage network failure, offering prognostic information not captured by molecular markers alone (Alexandersen et al., 2023; Nakamura et al., 2018).

Limitations include the relatively small sample size, reflecting the difficulty of acquiring longitudinal multimodal data in at-risk but asymptomatic individuals, and class imbalance (31 progressors vs. 71 non-progressors). While Cox models are robust to moderate class imbalance (Cox, 1972), the sample size precluded the use of cross-validation or more complex machine learning approaches. Biomarkers were not always collected on the same day, introducing modest timing offsets. However, sensitivity analyses using actual acquisition dates of plasma collection—rather than aligning them to MEG/MRI/PET session dates—yielded nearly identical results, supporting robustness.

In summary, our results demonstrate that integrating clinical, neurophysiological, and molecular biomarkers improves prediction of MCI progression in asymptomatic individuals. Importantly, the prognostic value of both alpha-band activity and neocortical Aβ PET evolves over time. These findings highlight the importance of modeling temporal dynamics in predictive frameworks for AD and position neurophysiological measures as valuable functional correlates of disease progression.

## Methods

### Participants

We analyzed data from a subset of participants from the PRe-symptomatic EValuation of Experimental or Novel Treatments for Alzheimer’s Disease (PREVENT-AD) cohort, a longitudinal study of cognitively unimpaired older adults with elevated familial risk for sporadic AD (Breitner et al., 2016). Elevated risk was defined as having one parent or multiple siblings diagnosed with AD (Breitner et al., 2016; Tremblay-Mercier et al., 2021). Inclusion criteria required a Montreal Cognitive Assessment score ≥ 26 (Nasreddine et al., 2005) and Clinical Dementia Rating of 0 (Hughes et al., 1982).

The main analytic sample comprised 124 participants who underwent Aβ and tau PET imaging and resting-state MEG between 2017 and 2019. All were cognitively unimpaired at biomarker collection (see *Neuropsychological assessment and MCI diagnosis* section below). We excluded 19 participants due to high-amplitude MEG noise from ferromagnetic dental implants, one for poor-quality PET data, and two with missing plasma measurements, yielding 102 participants for analysis. Group characteristics for progressors (*n* = 31) and non-progressors (*n* = 71) are provided in **Table 1**. Group differences were assessed using independent-sample Wilcoxon rank-sum tests for continuous variables and chi-square tests for categorical variables. Extended analyses included larger PREVENT-AD subsamples with plasma or PET biomarkers but no MEG: 211 participants with plasma data, 226 with Aβ PET, and 224 with tau PET.

All procedures were approved by the McGill Institutional Review Board and/or the Douglas Mental Health University Institute Research Ethics Board. All participants gave written informed consent.

### Neuropsychological assessment and MCI diagnosis

PREVENT-AD participants complete annual cognitive testing with the Repeatable Battery for the Assessment of Neuropsychological Status (RBANS; Randolph et al., 1998), which evaluates immediate memory, delayed memory, attention, visuospatial constructional abilities, and language. Decline in at least one domain prompted review by a multidisciplinary consensus panel at the Centre for Studies on Prevention of Alzheimer’s Disease (StoP-AD). The panel, blinded to *APOE* genotype and all biomarker data, determined MCI status, based on longitudinal neuropsychological, clinical, and medical information. Among the 102 participants in the main sample 31 progressed to MCI, with a mean time from biomarker collection to diagnosis of 4.04 years (SD = 1.90; range 0.19—7.02 years).

### APOE ε4 carrier status genotyping

Genomic DNA was extracted from whole blood (200 μl) using the QIASymphony system and the DNA Blood Mini QIA Kit (Qiagen, Valencia, CA, USA). *APOE* genotype was determined by pyrosequencing (PyroMark) of rs429358 and rs7412 polymorphisms, classifying participants as ε4 carriers or non-carriers.

### Structural magnetic resonance imaging

T1-weighted MRI data was acquired on a 3T Siemens TIM Trio scanner (12-or 32-channel head coil; Siemens Medical Solutions, Munich, Germany) at the Brain Imaging Center of the Douglas Mental Health University Institute (Montreal, Quebec, Canada). The magnetization-prepared rapid acquisition gradient echo (MPRAGE) sequence used the following parameters: 176 sagittal slices, 1-mm slice thickness, TR = 2300 ms, TE = 2.98 ms, flip angle = 9°, FOV = 256 × 240 × 176 mm. The scan closest to MEG/PET acquisition was selected (mean interval = 0.84 years; SD = 0.63). Cortical surfaces were reconstructed with *FreeSurfer* v5.3 (Fischl, 2012) and parcellated according to the Desikan-Killiany atlas (Desikan et al., 2006). This parcellation was used for both MEG source mapping and the regional quantification of Aβ and tau PET uptake. Hippocampal volumes were normalized to the total intracranial volume (TIV), averaged bilaterally, and expressed as percentage of brain volume.

### Magnetoencephalography

Two 5-minute resting-state (eyes-open) runs were recorded on a 275-channel CTF MEG system (Port Coquitlam, British Columbia, Canada). Data were sampled at 2400 Hz, with anti-aliasing (600 Hz). Environmental noise was reduced using third-order gradient compensation. Head position was tracked continuously. ECG/EOG were recorded for artifact correction, and empty-room data were collected before each session. MEG was acquired on the same day as either the Aβ or tau PET scan for most participants (mean interval from Aβ PET = 0.06 years; SD = 0.19).

Preprocessing followed *Brainstorm*’s protocols (Niso et al., 2019; Tadel et al., 2011) and good-practice guidelines (Gross et al., 2013): notch filters at 60 Hz harmonics; 0.3 Hz high-pass filter; bad channels/segments removed; signal space projectors (SSPs) computed from ECG/EOG; segmentation into 4-second epochs; semi-automated artifact rejection.

Source activity was reconstructed with depth-weighted dynamic statistical parametric mapping (dSPM) at 15,000 cortical vertices, using individual MRI-derived head models and constraining currents perpendicular to the cortical surface. PSDs were estimated per epoch (Welch’s method; 2-second windows, 50% overlap), averaged, and normalized to total power. Relative alpha-band (8–12 Hz) power was averaged across six bilateral temporal-lobe regions (entorhinal, fusiform, inferior parietal, lingual, middle temporal, parahippocampal) and used as the MEG biomarker (Niso et al., 2019).

### Plasma biomarkers

Blood was collected annually from PREVENT-AD participants as part of the study protocol. For this analysis, we selected the plasma sample obtained closest in time to the MEG/PET imaging session (mean difference = 0.91 years; SD = 0.89 years). Two plasma biomarkers were evaluated: 1) the Aβ42/40 ratio, which reflects levels of toxic soluble Aβ species in circulation and indexes Aβ metabolism (Blennow et al., 2015). Lower ratios are associated with increased amyloid plaque burden in the brain (Doecke et al., 2020; Fandos et al., 2017); and 2) phosphorylated tau at threonine 217 (p-tau217), a marker of both tau phosphorylation and amyloid pathology (Ashton et al., 2024; Gonzalez-Ortiz et al., 2023; Jonaitis et al., 2023).

The Aβ42/40 ratio was derived by quantifying plasma Aβ 1-42 and Aβ 1-40 concentrations using immunoprecipitation mass spectrometry (MS). Aβ peptides were immunoprecipitated with specific antibodies on a KingFisher Flex system and analyzed by liquid chromatography-tandem mass spectrometry (LC-MS/MS; Meyer et al., 2022). Plasma p-tau217 levels were measured using an ultrasensitive in-house single molecule array (Simoa) assay developed at the University of Gothenburg on the HD-X platform (UGOT p-tau217; Gonzalez-Ortiz et al., 2023), with an overall intraplate variation and intermediate precision < 10%.

In addition to the 102 participants in the main analysis, a further 109 PREVENT-AD participants had plasma biomarker data, yielding a total of 211 individuals in the extended plasma dataset (progressors = 48, mean time from biomarker collection to diagnosis = 4.93 years, SD = 2.01 years; range 1.02—10.45 years). These data were used to replicate the models incorporating clinical information and plasma biomarkers (**Supplementary Figure 2; Supplementary Figure 5**).

### Aβ and tau positron emission tomography

Aβ and tau PET imaging data were acquired and processed as described by Gallego-Rudolf et al., (2024). Scans were performed using a Siemens high-resolution research tomograph (HRRT) PET camera at the McConnell Brain Imaging Center, Montreal Neurological Institute. Aβ PET scans were obtained 40 to 70 minutes after intravenous injection of ∼6 mCi of [18F] NAV4694; tau PET scans were acquired 80 to 100 minutes following injection of ∼10 mCi [18F] AV-1451 (FTP). For most participants (*n* = 94), the two PET scans were acquired on consecutive days.

In addition to the main sample, 124 participants had available Aβ PET data and 122 had available tau PET data, resulting in extended samples of 226 individuals for Aβ PET (progressors = 53; mean time from biomarker collection to diagnosis = 3.31 years, SD = 2.06; range 0.06—7.02 years) and 224 for tau PET (progressors = 53; mean time from biomarker collection to diagnosis of 3.29 years, SD = 2.04; range 0.06—7.02 years). These extended samples were used to replicate the Cox regression models combining PET biomarkers with clinical information (**Supplementary Figure 2; Supplementary Figure 5**).

PET images were preprocessed using an in-house pipeline developed by the Villeneuve lab (https://github.com/villeneuvelab/vlpp). The 4D dynamic image files (6 × 5-minute frames for NAV; 4 × 5-minute frames for FTP) were realigned, averaged, and co-registered to each participant’s structural MRI. Standardized uptake value ratio (SUVR) maps were computed using the cerebellar gray matter as the reference for Aβ scans and the inferior cerebellar gray matter for tau scans. SUVR values were averaged across Desikan-Killiany atlas regions. Neocortical Aβ uptake was summarized as a size-weighted mean SUVR across early Aβ-accumulating regions (precuneus, posterior cingulate, parietal, frontal, and lateral temporal; Palmqvist et al., 2017; Villeneuve et al., 2015). Entorhinal tau PET uptake was selected as the primary tau feature due to its early involvement in AD pathology (Braak & Braak, 1991, 1995).

### Cox regression proportional hazard models

To assess the added value of integrating neurophysiological activity and proteinopathy biomarkers with clinical information for predicting progression to MCI, we fitted Cox regression proportional hazard models using the *survival* package in *R* (Therneau, 2024). The reference clinical model (Model 1) included demographic and genetic covariates: age at MEG acquisition, sex, years of education, and *APOE* ε4 carrier status (binary).

Subsequent models added individual biomarker measures: Model 2: MEG-derived alpha-band relative power (temporal lobe regions); Model 3: MRI-derived hippocampal volume (normalized to the total intracranial volume); Model 4: Plasma Aβ42/40 ratio and p-tau217; Model 5: Neocortical Aβ PET SUVR; Model 6: Entorhinal tau PET SUVR; Model 7: All available biomarkers combined.

Models were fitted with *coxph;* proportional hazards assumptions were tested using *cox.zph*. Survival curves were generated with *survminer* (Kassambara et al., 2024). Model performance was assessed via likelihood ratio tests (LRTs) against the null model (FDR-corrected, *n* = 7). Pairwise LRTs compared each biomarker model to the clinical model (FDR-corrected, *n* = 6). We also computed ΔAIC values (ΔAIC > 2 indicating preference for the model with a lower AIC; Burnham & Anderson, 1998) and the concordance index (C-index) for assessing discriminative accuracy (see **Supplementary Figure 1**). The C-index reflects how model predictions align with observed outcomes, with values closer to 1 indicating better performance. Pearson correlations among continuous biomarkers were computed to assess collinearity (**Supplementary Figure 3**; FDR-corrected, *n* = 28).

To visualize discriminative accuracy, participants were ranked by model-derived risk scores and split into tertiles (n = 34) representing low-, medium-, and high-risk groups; survival curves were then plotted. A greater separation between the survival curves indicates better model discrimination.

We assessed the robustness of our findings by replicating the Cox regression analyses using the extended sample of participants with available plasma and PET data (**Supplementary Figure 2**).

Within the full model, we assessed associations between biomarkers and risk scores using linear regression for continuous predictors and t-tests for categorical predictors (sex, *APOE* status), with FDR correction (*n* = 10). This analysis was repeated for the clinical model and each biomarker model (see **Supplementary Table 4**), applying FDR correction based on the number of predictors included in each model.

For time-dependent effects, we added biomarker × time interaction terms to each of the models combining clinical and biomarker information, entering them individually while keeping the rest of the predictors as linear terms. Hazard ratio trajectories were plotted from main and interaction coefficients, and time-varying risk associations were visualized across follow-up years, while accounting for clinical covariates. Replications were performed in extended samples (**Figure 4; Supplementary Figure 4; Supplementary Figure 5**). This approach enabled visualization of how each biomarker’s contribution to risk evolved over time.

We further tested whether including time-varying alpha power improved prediction beyond proteinopathy biomarkers by computing LRTs comparing clinical + plasma, clinical + Aβ PET, and clinical + tau PET models with and without the alpha × time term. A stepwise selection procedure (*step*, base *R*) was applied to the full model (all biomarkers + significant time interactions) to identify a parsimonious predictor set.

All statistical analyses were conducted in *R* v4.1.1 (R Core Team, 2021). Survival models were fitted using the *survival R* package (v3.5.0; Therneau, 2024). Stepwise regression used the base *R step* function. Figures and visualizations were generated using *ggplot2* (v3.3.5; Wickham, 2016), *ggseg* (v1.6.5; Mowinckel & Vidal-Piñeiro, 2020), *survminer* (v0.5.0; Kassambara et al., 2024), and *corrplot* (v0.92; Wei & Simko, 2024).

## Data availability

Data used in the preparation of this manuscript were obtained from the Pre-symptomatic Evaluation of Experimental or Novel Treatments for Alzheimer’s Disease (PREVENT-AD, https://prevent-alzheimer.net/). Some data are publicly available through the following platforms: (https://openpreventad.loris.ca/, and https://www.mcgill.ca/bic/neuroinformatics/omega (Niso et al., 2016). In compliance with the ethical and privacy policies stipulated in the PREVENT-AD study—which protect participant identity and allow participants the right to refrain to withhold sharing certain biological/imaging data—the full internal release of the dataset (https://registeredpreventad.loris.ca/) can only be shared upon approval by the scientific committee at the Centre for Studies on Prevention of Alzheimer’s Disease (StoP-AD) at the Douglas Research Centre. Detailed instructions for accessing both the open and full internal release versions of the PREVENT-AD dataset are available here: https://prevent-alzheimer.net/?page_id=1760&lang=en. For more information regarding the organization of the PREVENT-AD dataset, please refer to Breitner et al., 2016 and Tremblay-Mercier et al., 2021, or contact corresponding author Sylvia Villeneuve. The Desikan-Killiany atlas parcellation is included in *FreeSurfer* v5.3 (https://surfer.nmr.mgh.harvard.edu/fswiki/CorticalParcellation) and the *ggseg* package v1.6.5 (https://github.com/ggseg).

## Code availability

MRI parcellation was performed using *FreeSurfer* (v5.3; Fischl, 2012). PET analysis utilized a standard pipeline available at https://github.com/villeneuvelab/vlpp. The 3D OSEM algorithm used for PET imaging reconstruction has been described in previous publications (Sibomana et al., 2012; Varrone et al., 2009). MEG data analysis was conducted using *Brainstorm3* (Niso et al., 2019; Tadel et al., 2011), running on *Matlab* (vR2021b; The MathWorks Inc., 2021). All statistical analysis reported in this manuscript were performed using standard functions in *R* (v4.1.1; R Core Team, 2021). Cox proportional hazard regression models were implemented using the *cox.ph* function from R’s *survival* package (v3.5.0; Therneau, 2024). Data visualization was conducted with *ggplot2* (v3.4.0; Wickham, 2016) using *survminer* (v0.5.0; Kassambara et al., 2024) for plotting the survival curves and *ggseg* (v1.6.5; Mowinckel & Vidal-Piñeiro, 2020) for generating the brain plots. Results from the correlation analyses were visualized with the *corrplot R* package (v0.92; Wei & Simko, 2024). The scripts used to implement the statistical analyses presented in this manuscript are available in an open-access GitHub repository: https://github.com/jogaru1818/progression_to_MCI.

## Data Availability

Data used in the preparation of this manuscript were obtained from the Pre-symptomatic Evaluation of Experimental or Novel Treatments for Alzheimer’s Disease (PREVENT-AD, https://prevent-alzheimer.net/). Some data are publicly available through the following platforms: (https://openpreventad.loris.ca/, and https://www.mcgill.ca/bic/neuroinformatics/omega; Niso et al., 2016). In compliance with the ethical and privacy policies stipulated in the PREVENT-AD study—which protect participant identity and allow participants the right to refrain to withhold sharing certain biological/imaging data—the full internal release of the dataset (https://registeredpreventad.loris.ca/) can only be shared upon approval by the scientific committee at the Centre for Studies on Prevention of Alzheimer’s Disease (StoP-AD) at the Douglas Research Centre. Detailed instructions for accessing both the open and full internal release versions of the PREVENT-AD dataset are available here: https://prevent-alzheimer.net/?page_id=1760&lang=en. For more information regarding the organization of the PREVENT-AD dataset, please refer to Breitner et al., 2016 and Tremblay-Mercier et al., 2021, or contact corresponding author Sylvia Villeneuve. The Desikan-Killiany atlas parcellation is included in FreeSurfer v5.3 (https://surfer.nmr.mgh.harvard.edu/fswiki/CorticalParcellation) and the ggseg package v1.6.5 (https://github.com/ggseg).

https://openpreventad.loris.ca/

https://www.mcgill.ca/bic/neuroinformatics/omega

https://registeredpreventad.loris.ca/

## Acknowledgements

Data used in the preparation of this article were obtained from the PRe-symptomatic EValuation of Experimental or Novel Treatments for Alzheimer’s Disease (PREVENT-AD) cohort. The PREVENT-AD was launched in 2011 as a $13.5 million, 7-year public-private partnership using funds provided by McGill University. It was further funded by the Fonds de Recherche du Québec – Santé (FRQ-356162), an unrestricted research grant from Pfizer Canada, the J.L. Levesque Foundation, the Douglas Hospital Research Centre and Foundation, the Government of Canada, the Canada Fund for Innovation, the Canadian Institutes of Health Research (178385, 162091, 148963, 153287, 178210, 165921, 175328), the National Institutes of Health of the United States (NIH AG068563) and the Brain Canada Foundation. The authors acknowledge all the PREVENT-AD participants and their families, the PREVENT-AD team members, the Brain Imaging Center at the Douglas Research Centre and the MEG, PET and cyclotron units at the Montreal Neurological Institute for their time and dedication to this project. A complete list of PREVENT-AD acknowledgements can be found at: https://preventad.loris.ca/acknowledgements/acknowledgements.php?date=2025-09-17. The investigators of the PREVENT-AD program contributed to the design and implementation of PREVENT-AD and/or provided data but did not participate in the analysis or writing of this report. We also would like to thank the reviewers for providing their valuable insight, as well as Valentin Ourry, Mohammadali Javanray, Ting Qiu, Jordana Remz, Alfonso Fajardo-Valdez, Alexandre Pastor-Bernier and Frédéric St-Onge for providing advice on the study design. J.G.R. is supported by the Mexican National Council of Science and Technology (CONACyT; 2020-000017-02EXTF-00402) and funding from the Canada Research Excellence Fund and Fonds de recherche du Quebec through the Healthy Brains Healthy Lives (HBHL) initiative at McGill University. A.I.W. is supported by grant F32-NS119375 from the United States National Institutes of Health, a Banting Postdoctoral Fellowship (BPF-186555) from the Canadian Institutes of Health Research, and the Tier-2 Canada Research Chair in Neurophysiology of Aging and Neurodegeneration (CRC-2023-00300). Y.Y. is supported by the Alzheimer’s Society of Canada and the Fonds de Recherche du Québec – Santé. H.Z. is a Wallenberg Scholar and a Distinguished Professor at the Swedish Research Council, supported by grants from the Swedish Research Council (#2023-00356, #2022-01018 and #2019-02397), the European Union’s Horizon Europe research and innovation programme under grant agreement No 101053962, Swedish State Support for Clinical Research (#ALFGBG-71320), the Alzheimer Drug Discovery Foundation (ADDF), USA (#201809-2016862), the AD Strategic Fund and the Alzheimer’s Association (#ADSF-21-831376-C, #ADSF-21-831381-C, #ADSF-21-831377-C, and #ADSF-24-1284328-C), the European Partnership on Metrology, co-financed from the European Union’s Horizon Europe Research and Innovation Programme and by the Participating States (NEuroBioStand, #22HLT07), the Bluefield Project, Cure Alzheimer’s Fund, the Olav Thon Foundation, the Erling-Persson Family Foundation, Familjen Rönströms Stiftelse, Familjen Beiglers Stiftelse, Stiftelsen för Gamla Tjänarinnor, Hjärnfonden, Sweden (#FO2022-0270), the European Union’s Horizon 2020 research and innovation programme under the Marie Skłodowska-Curie grant agreement No 860197 (MIRIADE), the European Union Joint Programme – Neurodegenerative Disease Research (JPND2021-00694), the National Institute for Health and Care Research University College London Hospitals Biomedical Research Centre, the UK Dementia Research Institute at UCL (UKDRI-1003), and an anonymous donor. K.B. is supported by the Swedish Research Council (#2017-00915 and #2022-00732), the Swedish Alzheimer Foundation (#AF-930351, #AF-939721, #AF-968270, and #AF-994551), Hjärnfonden, Sweden (#ALZ2022-0006, #FO2024-0048-TK-130 and FO2024-0048-HK-24), the Swedish State under the agreement between the Swedish government and the County Councils, the ALF-agreement (#ALFGBG-965240 and #ALFGBG-1006418), the European Union Joint Program for Neurodegenerative Disorders (JPND2019-466-236), the Alzheimer’s Association 2021 Zenith Award (ZEN-21-848495), the Alzheimer’s Association 2022-2025 Grant (SG-23-1038904 QC), La Fondation Recherche Alzheimer (FRA), Paris, France, the Kirsten and Freddy Johansen Foundation, Copenhagen, Denmark, Familjen Rönströms Stiftelse, Stockholm, Sweden, and an anonymous philanthropist and donor. S.B. is supported by the United States National Institutes of Health (NIH, R01-EB026299-05), the Tier-1 Canadian Institutes of Health Research Canada Research Chair of Neural Dynamics of Brain Systems (CRC-2017-00311), a Discovery Grant from the Natural Sciences and Engineering Research Council of Canada (436355-13), the Canada Brain Research Fund (CBRF), an innovative arrangement between the Government of Canada (through Health Canada) and Brain Canada Foundation and the Alzheimer’s Association (AARG-NTF 926696). S.V. is supported by the Alzheimer Society of Canada, the Alzheimer’s Association, the Tier-1 Canada Research Chair in Early Detection of Alzheimer’s Disease and the Canadian Institutes of Health Research (CIHR, 178385, 162091, 148963).

## Author contributions statement

J.G.R., A.I.W., S.B. and S.V. conceptualized the study and wrote the manuscript, J.G.R., A.I.W., Y.Y., H.Z., K.B., S.B. and S.V. reviewed and edited the manuscript and were responsible for methodology. J.G.R. handled software development and conducted formal analysis. J.G.R., A.I.W. and Y.Y. validated the data. J.G.R., Y.Y., S.B. and S.V. performed data curation. J.G.R. handled study visualization. J.G.R., S.B. and S.V. led the investigation. S.B., S.V. contributed to supervision and project administration. H.Z., K.B., S.B. and S.V. contributed to resources and funding acquisition. A complete list of contributors to the PREVENT-AD Research Group is available at: https://preventad.loris.ca/acknowledgements/acknowledgements.php?DR=8.0&authors.

## Competing interest statement

H.Z. has served at scientific advisory boards and/or as a consultant for Abbvie, Acumen, Alector, Alzinova, ALZpath, Amylyx, Annexon, Apellis, Artery Therapeutics, AZTherapies, Cognito Therapeutics, CogRx, Denali, Eisai, Enigma, LabCorp, Merck Sharp & Dohme, Merry Life, Nervgen, Novo Nordisk, Optoceutics, Passage Bio, Pinteon Therapeutics, Prothena, Quanterix, Red Abbey Labs, reMYND, Roche, Samumed, ScandiBio Therapeutics AB, Siemens Healthineers, Triplet Therapeutics, and Wave, has given lectures sponsored by Alzecure, BioArctic, Biogen, Cellectricon, Fujirebio, LabCorp, Lilly, Novo Nordisk, Oy Medix Biochemica AB, Roche, and WebMD, is a co-founder of Brain Biomarker Solutions in Gothenburg AB (BBS), which is a part of the GU Ventures Incubator Program, and is a shareholder of MicThera (the work presented in this paper). KB has served as a consultant and at advisory boards for Abbvie, AC Immune, ALZPath, AriBio, Beckman-Coulter, BioArctic, Biogen, Eisai, Lilly, Moleac Pte. Ltd, Neurimmune, Novartis, Ono Pharma, Prothena, Quanterix, Roche Diagnostics, Sunbird Bio, Sanofi and Siemens Healthineers; has served at data monitoring committees for Julius Clinical and Novartis; has given lectures, produced educational materials and participated in educational programs for AC Immune, Biogen, Celdara Medical, Eisai and Roche Diagnostics and is a co-founder of Brain Biomarker Solutions in Gothenburg AB (BBS), which is a part of the GU Ventures Incubator Program, outside the work presented in this paper. J.G.R., A.I.W., Y.Y., S.B. and S.V. declare no competing interests.

## Notes

### Author Declarations

The Institutional Review Boards of McGill University and the Douglas Mental Health University Institute gave ethical approval for this work.

