## Supplementary Material for "Prediction of mild cognitive impairment progression using time-sensitive multimodal biomarkers"

| Biomarker | Non-progressors ($n$ = 163) | Progressors to MCI ($n$ = 48) | Group differences |
|---|---|---|---|
| **Clinical** | | | |
| Age (years) | 64.87 (5.41) | 67.66 (4.67) | **$W$ = 2617.5; $p_{(FDR)}$ = 0.0010** |
| Sex female (%) | 118 (72.39) | 37 (77.08) | $\chi^2$ = 0.21; $p_{(FDR)}$ = 0.6448 |
| Education (years) | 15.40 (3.10) | 14.12 (3.21) | **$W$ = 4743; $p_{(FDR)}$ = 0.0370** |
| *APOE* ε4 carriers (%) | 57 (34.96) | 25 (52.08) | $\chi^2$ = 3.87; $p_{(FDR)}$ = 0.0587 |
| **Plasma** | | | |
| Aβ42/40 ratio (no units) | 0.096 (0.014) | 0.088 (0.014) | **$W$ = 5236.5; $p_{(FDR)}$ = 0.0010** |
| p-tau217 (pg/ml) | 2.34 (1.47) | 3.42 (1.57) | **$W$ = 2100.5; $p_{(FDR)}$ < 0.00001** |

**Supplementary Table 1. Characteristics of the extended PREVENT-AD sample with plasma data.** Comparison of clinical and plasma biomarker measurements between individuals who remained cognitively unimpaired (non-progressors; $n$ = 163) and individuals who developed MCI (progressors; $n$ = 48) in the extended PREVENT-AD dataset comprising all participants with available plasma data ($n$ = 211). Continuous variables are reported as means (standard deviations) and categorical variables (sex and *APOE* ε4 carrier status) are presented as counts (percentages). Group differences were assessed using independent-sample Wilcoxon rank-sum tests for continuous variables and chi-square tests for categorical variables. Statistical significance was set at $p$-values < 0.05, and significant $p$-values are shown in boldface. Consistent with the main sample, progressors were significantly older, had lower plasma Aβ42/40 ratios, and higher plasma p-tau217 levels. In this extended sample, progressors also had fewer years of education. Pg/ml, picograms per milliliter.

| **Biomarker** | **Non-progressors ($n$ = 173)** | **Progressors to MCI ($n$ = 53)** | **Group differences** |
|---|---|---|---|
| **Clinical** | | | |
| Age (years) | 67.55 (4.91) | 69.28 (4.57) | $\boldsymbol{W = 3637; p_{(FDR)} = 0.0383}$ |
| Sex female (%) | 119 (68.78) | 40 (75.47) | $\chi^2 = 0.57; p_{(FDR)} = 0.4469$ |
| Education (years) | 15.73 (3.09) | 14.40 (3.19) | $\boldsymbol{W = 5576.5; p_{(FDR)} = 0.0383}$ |
| *APOE* ε4 carriers (%) | 66 (38.15) | 26 (49.05) | $\chi^2 = 1.57; p_{(FDR)} = 0.2622$ |
| **Aβ PET** | | | |
| Neocortical Aβ (SUVR) | 1.26 (0.22) | 1.49 (0.40) | $\boldsymbol{W = 3017.5; p_{(FDR)} = 0.0008}$ |

**Supplementary Table 2. Characteristics of the extended PREVENT-AD sample with Aβ PET data.** Comparison of clinical variables and neocortical Aβ PET uptake between individuals who remained cognitively unimpaired (non-progressors; $n$ = 173) and individuals who developed MCI (progressors; $n$ = 53) in the extended PREVENT-AD dataset comprising all participants with available Aβ PET scans ($n$ = 226). Continuous variables are reported as means (standard deviations) and categorical variables (sex and *APOE* ε4 carrier status) are presented as counts (percentages). Group differences were assessed using independent-sample Wilcoxon rank-sum tests for continuous variables and chi-square tests for categorical variables. Statistical significance was set at $p$-values < 0.05, and significant $p$-values are shown in boldface. Consistent with the main sample, progressors were significantly older and had higher neocortical Aβ PET uptake. In this extended sample, progressors also had fewer years of education. SUVR, standardized uptake value ratio.

| Biomarker | Non-progressors (*n* = 171) | Progressors to MCI (*n* = 53) | Group differences |
|---|---|---|---|
| **Clinical** | | | |
| Age (years) | 67.60 (4.90) | 69.30 (4.58) | **$W$ = 3619.5; $p_{(FDR)}$ = 0.0450** |
| Sex female (%) | 119 (69.59) | 40 (75.47) | $\chi^2$ = 0.42; $p_{(FDR)}$ = 0.5150 |
| Education (years) | 15.76 (3.09) | 14.40 (3.19) | **$W$ = 5538; $p_{(FDR)}$ = 0.0354** |
| *APOE* ε4 carriers (%) | 65 (38.01) | 26 (49.05) | $\chi^2$ = 1.61; $p_{(FDR)}$ = 0.2549 |
| **Tau PET** | | | |
| Entorhinal cortex (SUVR) | 1.04 (0.11) | 1.14 (0.14) | **$W$ = 2649; $p_{(FDR)}$ = 0.00002** |

**Supplementary Table 3. Characteristics of the extended PREVENT-AD sample with tau PET data.** Comparison of clinical variables and entorhinal tau PET uptake between individuals who remained cognitively unimpaired (non-progressors; *n* = 171) and individuals that developed MCI (progressors; *n* = 53) in the extended PREVENT-AD dataset comprising all participants with available tau PET scans (*n* = 224). Continuous variables are reported as means (standard deviations), and categorical variables (sex and *APOE* ε4 carrier status) are presented as counts (percentages). Group differences were assessed using independent-sample Wilcoxon rank-sum tests for continuous variables and chi-square tests for categorical variables. Statistical significance was set at *p*-values < 0.05, and significant *p*-values are shown in boldface. Consistent with the main sample, progressors were significantly older and had higher entorhinal tau PET uptake. In this extended sample, progressors also had fewer years of education. SUVR, standardized uptake value ratio.

| Biomarker | Clinical | MEG | MRI | Plasma | Aβ PET | Tau PET |
|---|---|---|---|---|---|---|
| Age | $t$ = 8.12; $p_{(FDR)}$ < 0.00001 | $t$ = 6.77; $p_{(FDR)}$ < 0.00001 | $t$ = 7.15; $p_{(FDR)}$ < 0.00001 | $t$ = 5.13; $p_{(FDR)}$ < 0.00001 | $t$ = 4.83; $p_{(FDR)}$ < 0.00001 | $t$ = 5.31; $p_{(FDR)}$ < 0.00001 |
| Sex | $t$ = -8.00; $p_{(FDR)}$ < 0.00001 | $t$ = -6.43; $p_{(FDR)}$ < 0.00001 | $t$ = -7.12 $p_{(FDR)}$ < 0.00001 | $t$ = -5.03; $p_{(FDR)}$ < 0.00001 | $t$ = -5.42; $p_{(FDR)}$ < 0.00001 | $t$ = -5.55; $p_{(FDR)}$ < 0.00001 |
| Education | $t$ = -1.27; $p_{(FDR)}$ = 0.2071 | $t$ = -0.68; $p_{(FDR)}$ = 0.4981 | $t$ = -0.95; $p_{(FDR)}$ = 0.4322 | $t$ = -1.50; $p_{(FDR)}$ = 0.1360 | $t$ = -0.44; $p_{(FDR)}$ = 0.6595 | $t$ = -0.01; $p_{(FDR)}$ = 0.9919 |
| *APOE* | $t$ = -2.64; $p_{(FDR)}$ = 0.0132 | $t$ = -2.61; $p_{(FDR)}$ = 0.0132 | $t$ = -2.39; $p_{(FDR)}$ = 0.0319 | $t$ = -2.47; $p_{(FDR)}$ = 0.0184 | $t$ = -2.06; $p_{(FDR)}$ = 0.0533 | $t$ = -1.68; $p_{(FDR)}$ = 0.1217 |
| Alpha power | NA | $t$ =5.82; $p_{(FDR)}$ < 0.00001 | NA | NA | NA | NA |
| Hipp. vol. | NA | NA | $t$ = 0.18 $p_{(FDR)}$ = 0.8540 | NA | NA | NA |
| Aβ42/40 ratio | NA | NA | NA | $t$ = -5.98; $p_{(FDR)}$ < 0.00001 | NA | NA |
| p-tau217 | NA | NA | NA | $t$ = 9.48; $p_{(FDR)}$ < 0.00001 | NA | NA |
| Aβ PET | NA | NA | NA | NA | $t$ = 15.98; $p_{(FDR)}$ < 0.00001 | NA |
| Tau PET | NA | NA | NA | NA | NA | $t$ = 16.16; $p$ < 0.00001 |

**Supplementary Table 4. Association between biomarker measurements and risk scores derived from each model.** Summary of statistical tests assessing the association between individual biomarker measurements and the risk scores estimated from the clinical model and from each model combining clinical information with a specific set of biomarkers. As in Figure 4, associations were tested using linear regression for continuous predictors (age, education, MEG alpha, hippocampal volume, plasma Aβ42/40 ratio, plasma p-tau217, Aβ PET, tau PET) and independent-sample *t*-tests for categorical predictors (sex and *APOE* ε4 status). For each model, we report the *t*-statistic and FDR-corrected *p*-value (adjusted for the number of features included in each model). "NA" indicates that a given biomarker was not included in that model. These results confirm that associations between individual biomarkers and model-derived risk scores are consistent across models and are not driven by collinearity. Hipp. vol., hippocampal volume.

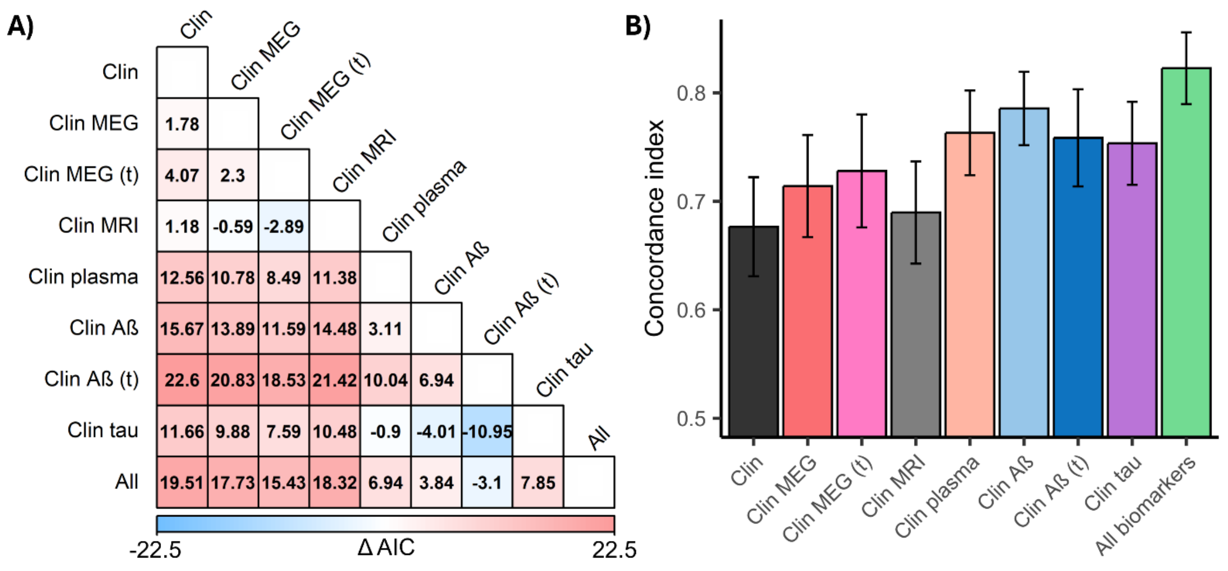


**Supplementary Figure 1. Comparison of Cox proportional hazard models. A)** Model performance comparison based on pairwise differences in Akaike Information Criterion (ΔAIC) values. A ΔAIC > 2 units is typically interpreted as evidence of better model fit for the model with the lower AIC value. Positive values (shaded red) indicate better performance of the model listed in the corresponding row; negative values (blue) indicate better performance of the model in the column. Models combining clinical information with plasma biomarkers, neocortical Aβ PET uptake, and entorhinal tau PET uptake outperformed the clinical model and the models adding MEG alpha power or hippocampal volume. Adding time-varying interaction terms for MEG alpha and Aβ PET (t) improved performance compared to their respective non-interaction models. The model including all biomarkers achieved the best performance overall. **B)** Bar plot of the concordance index (C-index) with standard error bars for each model. Higher C-index values reflect better discriminative accuracy. Results are consistent with the likelihood ratio tests and ΔAIC analyses, showing that models incorporating biomarker data—particularly plasma and PET proteinopathy—outperform the clinical model alone.

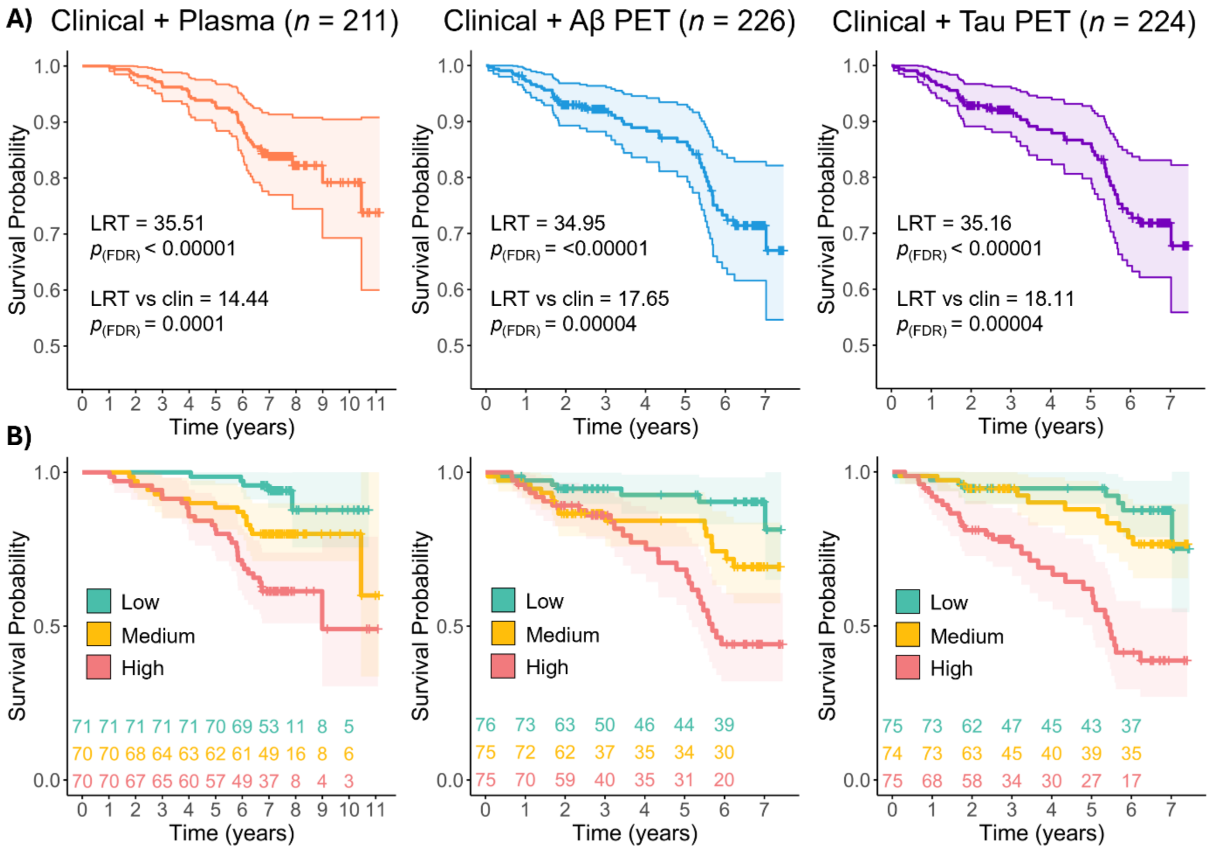


**Supplementary Figure 2. Replication of Cox regression models incorporating proteinopathy biomarkers in extended PREVENT-AD samples.** To assess the robustness of the main findings reported in Figure 2—namely, that proteinopathy biomarkers improve the prediction of progression to MCI—we replicated the Cox regression analyses in extended PREVENT-AD samples with available plasma ($n$ = 211), Aβ PET ($n$ = 226), and tau PET ($n$ = 224) data. **A)** Survival curves for each model combining clinical information with one set of proteinopathy biomarkers. Consistent with the main sample, likelihood ratio tests (LRTs) showed that adding plasma, Aβ PET, or tau PET biomarkers significantly improved prediction of MCI conversion compared with both the null and the clinical model alone (LRT vs clin). P-values were FDR-corrected to account for multiple comparisons. Thick lines represent estimated survival curves; shaded areas denote 95% confidence intervals. FDR, False discovery rate. **B)** Participants were stratified into low-, medium-, and high-risk tertiles based on model-derived risk scores, and survival curves were plotted for each group to illustrate model discriminative performance. Thick lines represent survival estimates per group; shaded areas show 95% confidence intervals. The number of cognitively stable participants remaining at each follow-up year is shown below each plot.

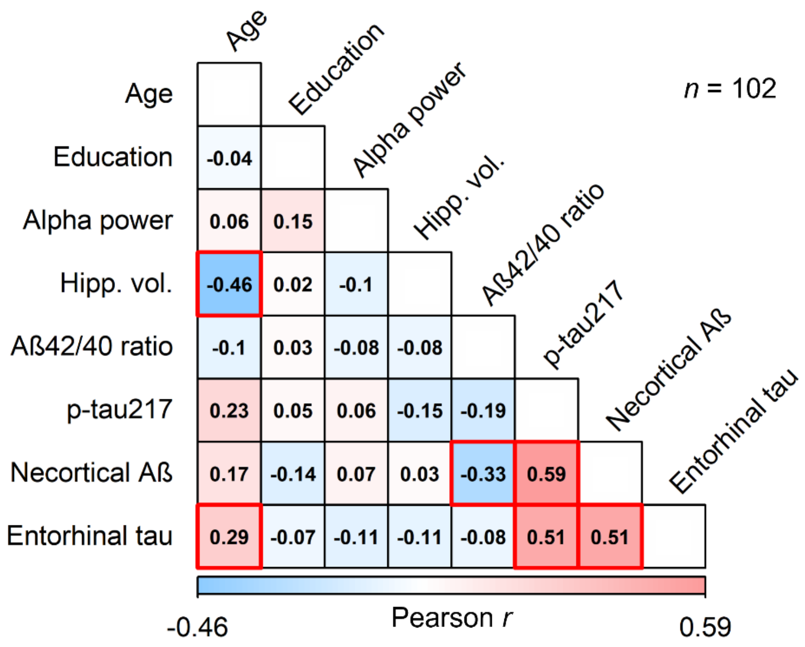


**Supplementary Figure 3. Assessment of potential collinearity among biomarkers in the full model.** Pairwise Pearson correlation coefficients (*r*) between all continuous variables included in the full Cox regression model that incorporated all biomarkers. Correlation strengths are shown using a blue-to-red color scale (blue for negative, red for positive) and annotated within each cell of the matrix. Red boxes highlight statistically significant correlations (FDR-corrected $p < 0.05$). Correlations values > 0.3 indicate moderate associations, while values above 0.5 indicate strong correlations. As expected, older age was significantly associated with lower hippocampal volume and higher entorhinal tau PET uptake, while plasma and PET measures of proteinopathy (Aβ42/40 ratio, p-tau217, neocortical Aβ, and entorhinal tau) were moderately to strongly correlated with each other. These results support the inclusion of all biomarkers in the full model while highlighting key interdependencies. Hipp. vol., hippocampal volume.

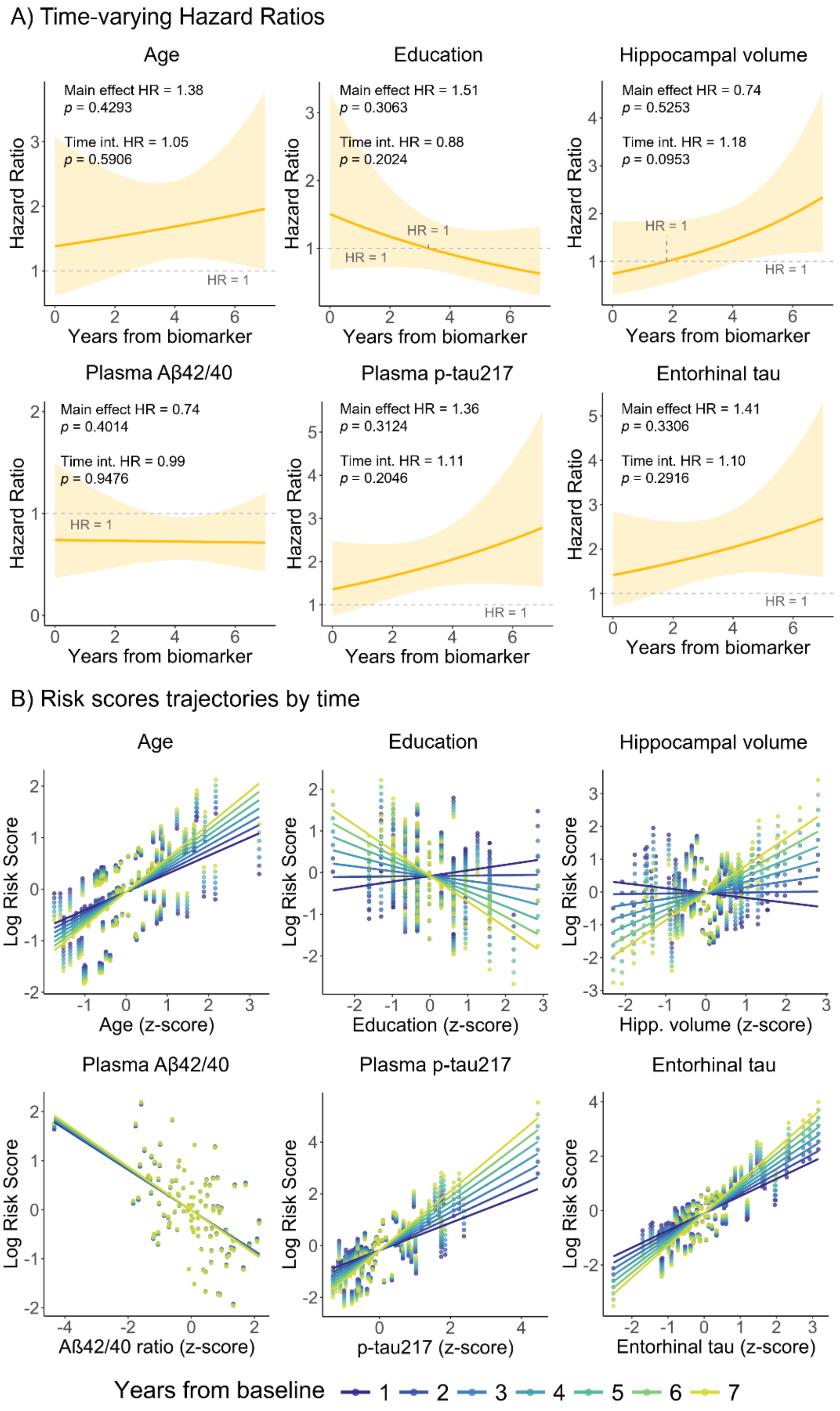

A) Time-varying Hazard Ratios
Age
Main effect HR = 1.38
p = 0.4293
Time int. HR = 1.05
p = 0.5906
HR = 1
Education
Main effect HR = 1.51
p = 0.3063
Time int. HR = 0.88
p = 0.2024
HR = 1
HR = 1
Hippocampal volume
Main effect HR = 0.74
p = 0.5253
Time int. HR = 1.18
p = 0.0953
HR = 1
HR = 1
Plasma Aβ42/40
Main effect HR = 0.74
p = 0.4014
Time int. HR = 0.99
p = 0.9476
HR = 1
Plasma p-tau217
Main effect HR = 1.36
p = 0.3124
Time int. HR = 1.11
p = 0.2046
HR = 1
Entorhinal tau
Main effect HR = 1.41
p = 0.3306
Time int. HR = 1.10
p = 0.2916
HR = 1
Hazard Ratio
Years from biomarker
B) Risk scores trajectories by time
Age
Education
Hippocampal volume
Plasma Aβ42/40
Plasma p-tau217
Entorhinal tau
Log Risk Score
Age (z-score)
Education (z-score)
Hipp. volume (z-score)
Aß42/40 ratio (z-score)
p-tau217 (z-score)
Entorhinal tau (z-score)
Years from baseline
1 2 3 4 5 6 7

**Supplementary Figure 4. Time-varying associations between biomarkers and risk of progression to MCI.** Cox regression models were fitted to include time-dependent interaction terms for each continuous biomarker in the clinical model and the models combining clinical information with each set of biomarkers. **A)** Hazard ratios over time, derived from the main effect and time interaction coefficients for each variable. None of the predictors (age, years of education, hippocampal volume, plasma Aβ42/40 ratio, plasma p-tau217, or entorhinal tau PET uptake) showed significant time-dependent changes in their associations with risk of MCI progression. Shaded areas represent 95% confidence intervals. The main effect and time interaction hazard ratio coefficients and associated *p*-values are embedded on each plot. HR, hazard ratio; Time int., time interaction. **B)** Time-varying associations between the biomarker values (z-scored) and predicted log-relative risk. Each line represents a different follow-up year (1–7), color-coded from dark blue (early) to yellow (late). Risk predictions account for all covariates in the model (age, sex, education, and *APOE* ε4 carrier status). Hipp. volume, hippocampal volume.

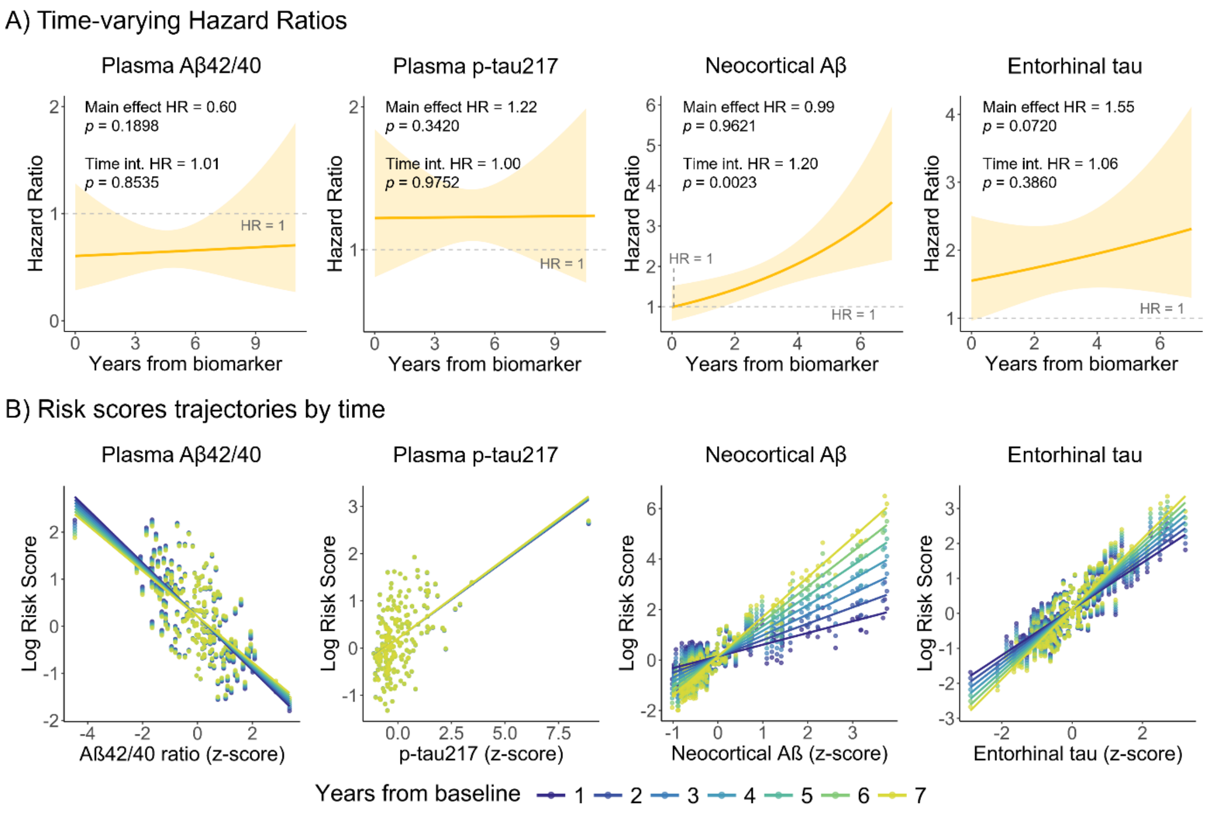


**Supplementary Figure 5. Replication of time-dependent Cox models for proteinopathy biomarkers in the extended samples.** We replicated our Cox regression models with time-dependent interaction terms for each proteinopathy biomarker using the extended sample of participants with available plasma (*n* = 211), Aβ PET (*n* = 226), and tau PET (*n* = 224) data. **A)** Hazard ratio trajectories over time, computed from the main effect and time interaction coefficients. Consistent with our primary analysis, the risk associated with neocortical Aβ PET uptake increased significantly with time. No significant time-dependent effects were found for plasma Aβ42/40 ratio, plasma p-tau217, or entorhinal tau PET uptake. The main effect and time interaction hazard ratio coefficients and associated *p*-values are embedded on each plot. HR, hazard ratio; Time int., time interaction. **B)** Time-varying associations between each z-scored biomarker and predicted log-relative risk. Each line corresponds to a follow-up year (1–7), color-coded from dark blue (early) to yellow (late). Risk predictions account for all covariates in the model (age, sex, education, *APOE* ε4 carrier status).
